# Factors associated with acceptance of COVID-19 vaccination among women in Guinea: Analysis of the first vaccination phase in March 2021

**DOI:** 10.1101/2023.03.27.23287835

**Authors:** Almamy Amara Touré, Ibrahima Barry, Aboubacar Sidiki Magassouba, Kadiatou Bah, Diao Cisse, Younoussa Sylla, Alsény Yarie Camara, Gaspard Loua, Abdourahamane Diallo

**Author notes:** These authors contributed equally to this work (Ibrahima Barry, Aboubacar Sidiki Magassouba). These authors also contributed equally to this work (Kadiatou Bah, Diao Cisse, Younoussa Sylla, Gaspard Loua, Abdourahamane Diallo).

## Abstract

Vaccination remains the primary strategy for ending the COVID-19 pandemic. However, vaccination rates are still low in low-income countries. The primary goal of this study was to describe the status of COVID-19 vaccine acceptance and hesitancy among women in Guinea and to identify associated predictors. *We* conducted a cross-sectional study in five Guinean cities (Conakry, Mamou, Kindia, Kankan and N’zérékoré) across the four natural regions between Mar 22 and Aug 25 2021. Participants aged 18 were randomly recruited from the healthcare workers (HCWs) and the general population (GP). We used multivariate logistic regression to identify facilitators and barriers to acceptance of COVID-19 vaccination and a classification and regression tree (CART) to extract the profile of predictors. *We included* 2,208 women among the HCWs and 1,121 in the GP. Most HCWs (63%) were already vaccinated, compared to only 28% of GP. The main factors associated with acceptance of a COVID-19 vaccine in the HCWs were an absence of pregnancy ORA = 4.46 [CI95%: 3.08, 6.52] and positive subjective norms ORA = 2.34 [CI95%: 1.92, 2.84].

Regarding the GP, the main factors were the ability to receive the vaccine ORA = 5.20 [CI95%: 3.45, 8.01] and being adult ORA = 2.25 [CI95%: 1.34, 3.79] associated with acceptance of vaccination. Vaccination rates were higher in the HCWs. Favourable subjective norms and ability to receive the vaccine were facilitators of acceptance of COVID-19 vaccination, while youth and pregnancy were barriers to the approval of the COVID-19 vaccine.

## 1 Introduction

The world faced ineffective treatment at the beginning of the COVID-19 pandemic [1,2]. Many interventions have played an essential role in controlling the spread of COVID-19 infection, including wearing a mask, quarantine, and social distancing [3,4]; these measures damage the economy and increase the lack of social ties, negatively impacting the physical and mental health of populations [5]. A previous study on the psychosocial impacts of COVID-19 in Guinea found that 54% of participants had lost their jobs [6]. Therefore, maintaining these restrictive interventions was not feasible in the long term [7]. SARS-CoV-2 vaccination remains the primary strategy to end the COVID-19 pandemic by establishing herd immunity in the general population [2,6,8].

However, low vaccination intention has been reported in women [9]. This state of fact could be problematic because the risk of contracting COVID-19 is higher among them [10]. Indeed, they represent 70% of the global health and social care workforce and are more likely to be the primary caregivers of sick parents [10].

The accelerated development process of COVID-19 vaccines [11,12] and misinformation regarding the benefits, drug composition, and adverse effects of these vaccines have limited overall adherence [13–15]. The high availability of vaccination doses is a necessary but insufficient prerequisite for adequate vaccination coverage [9]. “Vaccine hesitancy, which refers to the delay in accepting or refusing available vaccination, is a common public problem in the application and promotion of various vaccines” [2]. The World Health Organization (WHO) has even listed it as one of the top ten threats to global health in 2019 [15]. In Guinea, vaccination began on Mar 5 2021 [16]. As of Apr 15 2021, 80,992 (0.6%) people had received at least one dose of the vaccine, including 25,032 (0.19%) for the second dose [16,17].

Guinea means “woman” in one of the primary local languages. Like the world’s health care, Women in Guinea are more significant among Frontline health workers. Additionally, women are essential in economic activities such as trade and agriculture. Therefore, they are a more substantial population at risk of COVID-19. The acceptance of COVID-19 by women is an achievement towards herd immunity; they stand for around 52% of the Guinea inhabitants [18].

This study aimed to describe the acceptance and hesitancy of the COVID-19 vaccine among women in Guinea and to identify the associated predictors. Our findings could help inform health authorities in developing specific vaccination strategies.

## 2 Methodology

### 2.1 Type and period of study

We conducted a cross-sectional study between Mar 22 and Aug 25 2021.

### 2.2 Study setting

We studied in four Guinean cities (Conakry, Mamou, Kankan and N’zérékoré). It has been carried out simultaneously as another great study [19] and used the same methodology and analytical approach.

### 2.3 Study population

We randomly recruited participants from healthcare facilities and workplaces (general population).

### 2.4 Selection criteria

#### 2.4.1 Inclusion criteria

- Given free and informed consent;
- Be at least 18 years old at the time of inclusion;
- Be available and able to express yourself.

#### 2.4.2 Non-inclusion criteria

- Refusal to participate in the investigation.

### 2.5 Sampling

#### 2.5.1 Healthcare workers (HCWs)

As a first step, we randomly selected health facilities in each of the four cities based on the list of health facilities. Next, we chose the health workforce that met our inclusion criteria.

#### 2.5.2 The general population (GP)

After identifying workplaces based on initial recruitment, we focused only on the women who were there and who met our inclusion criteria.

### 2.6 Study variables

#### 2.6.1 Dependent variable

This variable is the vaccination status. Each participant was asked whether they were vaccinated. The modalities of the answers were Yes or No.

#### 2.6.2 Independent variables

They included:

##### 2.6.2.1 Sociodemographic characteristics

- Residence: Conakry, Mamou, Kankan, and N’zérékoré.
- Age: expressed in completed years. Recoded as adults (over 40 years old) or young (40 years old and under).
- Marital status: married or single.
- Education: Second, university, high school.
- Profession
  - HCWs: nurse assistant, Laboratory technician, Physician, Medical support, Midwife Internship.
  - GP: Private employees, students, Civil servants, Freelance, Unemployed.
- Number of persons in the household: used to construct the variable
- Average monthly income: used to build the variable #**household income**#. This variable was the participant’s average monthly income.
- Household income
  - High: if the average income is≥ 2,000,000 GNF and the number of persons in the household is ≤ 10.
  - Low: if the average income is < GNF 2,000,000 and the number of persons in the household >10.
  - Intermediate: if other situation.
- Pregnancy: Yes or No.

##### 2.6.2.2 Medical history

- High blood pressure (hypertension): Yes or No.
- Overweight: Yes or No.
- Asthma: Yes or No.
- Allergic conditions: these were either sinusitis, rhinitis, or vaccine allergy. The answer was Yes or No.
- Other chronic diseases: Yes or No.

##### 2.6.2.3 Existing knowledge of vaccine

Healthcare providers: This was whether the participant had prior knowledge about vaccination. Items relied on the definition of vaccination, vaccine types, post-injection adverse events, and individual and herd immunity. The answer was Yes or No.

General population: This was whether the participant was aware of the general principle of vaccination. The answer was Yes or No.

##### 2.6.2.4 Search for information on COVID vaccines in the last three days

Were the participants asked the following question: Have you recently searched for information about COVID-19? The answer was Yes or No.

##### 2.6.2.5 Sources of information

These are the sources of information used by the participants to learn about the disease. These were: social networks, state radio, state television, private radio, private television, neighbourhood, word of mouth, and the NAHS website.

##### 2.6.2.6 Perception/fear of COVID-19

This section has three items rated from 1 to 5 (strongly disagree, disagree, neutral, agree, strongly agree).

| items | Affirmations | Scale/response |
| --- | --- | --- |
|  |  | 1. strongly disagree |
| – Perceived Susceptibility of the disease | • I am likely to get COVID-19 | 2. disagree |
|  | • I am at risk of COVID-19 | 3. neutral |
|  | • I may get COVID-19 | 4. agree |
|  |  | 5. I strongly agree |
| – Perceived seriousness of the disease | • I think that COVID-19 is a severe health problem | 1. strongly disagree |
|  |  | 2. disagree |
|  | • I believe that COVID-19 has negative consequences | 3. neutral |
|  | • I guess that COVID-19 is highly harmful | 4. agree |
|  |  | 5. I strongly agree |
| – Fear of COVID | • I am terrified of COVID-19 |  |
|  | • Thinking about COVID-19 makes me feel uncomfortable |  |
|  | • My palms got sweaty when I think about COVID-19 | 1. strongly disagree |
|  | • Watching the news about COVID-19 on social networks makes me nervous or anxious | 2. disagree |
|  |  | 3. neutral |
|  | • I cannot sleep because I am fearful of catching COVID-19 | 4. agree |
|  | • My heart rate increases or I have palpitations when I think about COVID-19 | 5. I strongly agree |

##### 2.6.2.7 Attitudes and beliefs

This section consists of two (2) items rated from 1 to 5 (strongly disagree, disagree, neutral, agree, strongly agree).

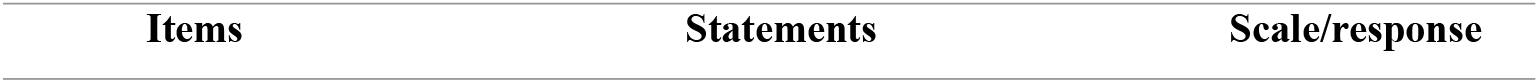
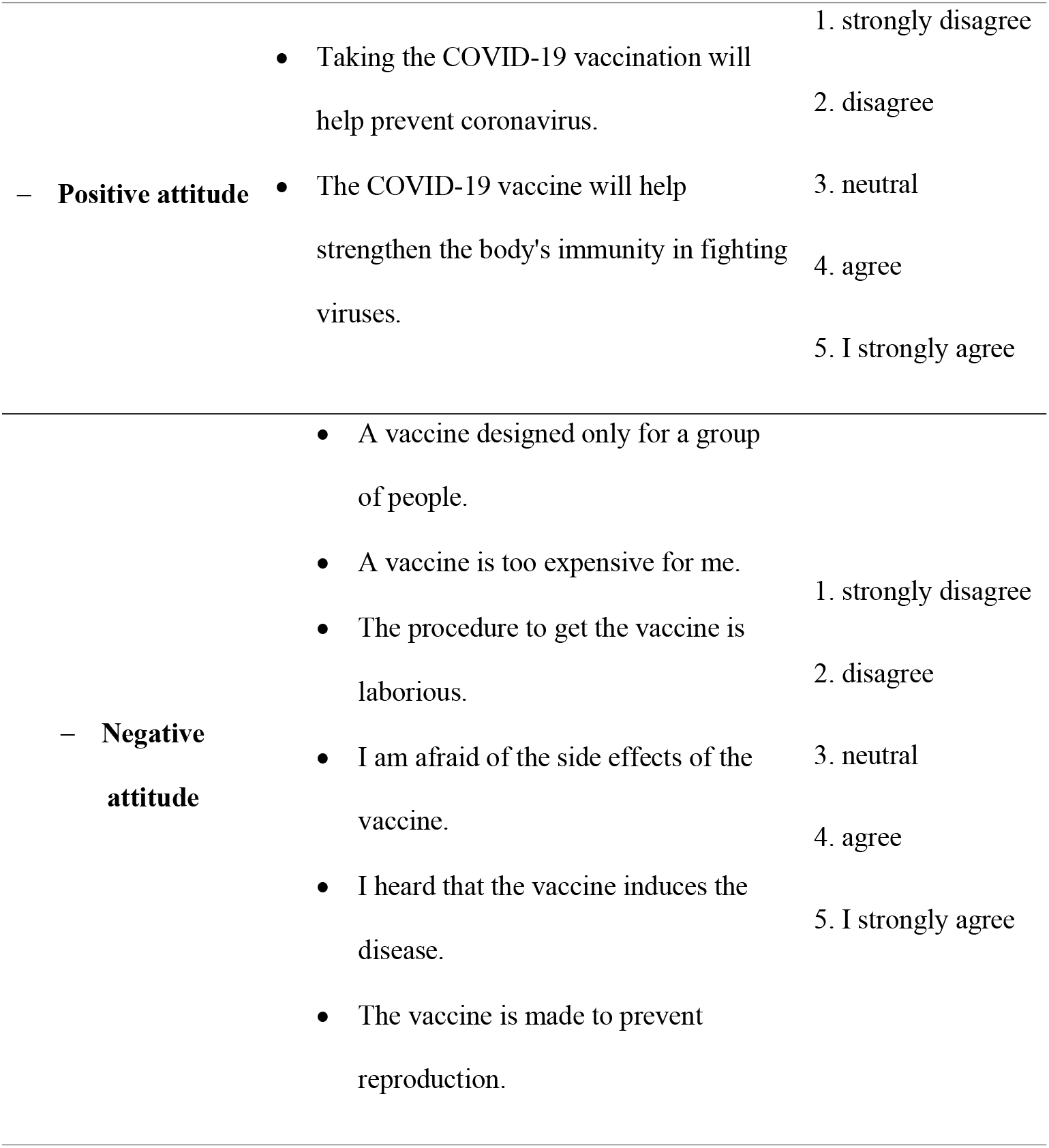

##### 2.6.2.8 Subjective norms

We asked the participants what they thought of the statements below. Responses were rated from 1 to 5 (strongly disagree, disagree, neutral, agree, strongly agree).

| Item | Statements | Scale/response |
| --- | --- | --- |
| **Subjective Norms** | • My parents and friends advised me to take the COVID-19 vaccine. | 1. strongly disagree |
|  | Many people in my neighbourhood think vaccination is an excellent way to prevent disease. | 2. disagree<br>3. neutral<br>4. agree |
|  | I have seen people like me getting vaccinated. | 5. I strongly agree |

##### 2.6.2.9 Ability to receive the vaccine

Evaluated the statements below and rated 1 to 5 (strongly disagree, disagree, neutral, agree, strongly agree).

2.6.2.9 Ability to receive the vaccine Evaluated the statements below and rated 1 to 5 (strongly disagree, disagree, neutral, agree, strongly agree).
| Item | Statements | Scale/response |
| --- | --- | --- |
|  |  | 1. strongly disagree |
|  | I can get the vaccine to avoid getting COVID-19 | 2. disagree |
| Ability | It is easy for me to get a vaccine to protect me from COVID-19 | 3. neutral<br>4. agree<br>5. strongly agree |

##### 2.6.2.10 Intent to receive COVID-19 vaccine

Evaluated the statements below and rated 1 to 5 (strongly disagree, disagree, neutral, agree, strongly agree).

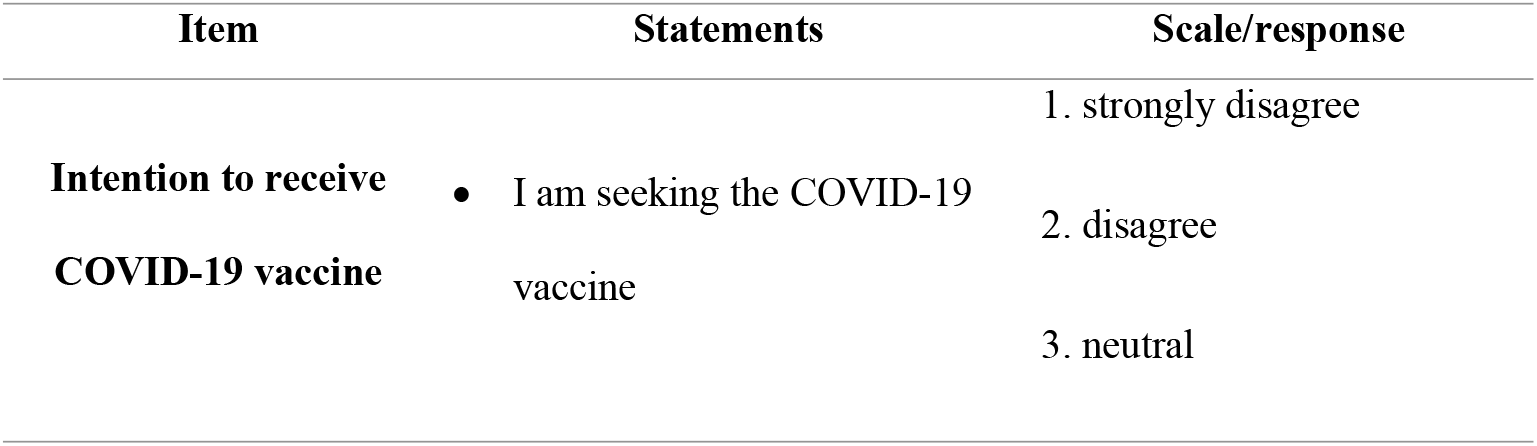
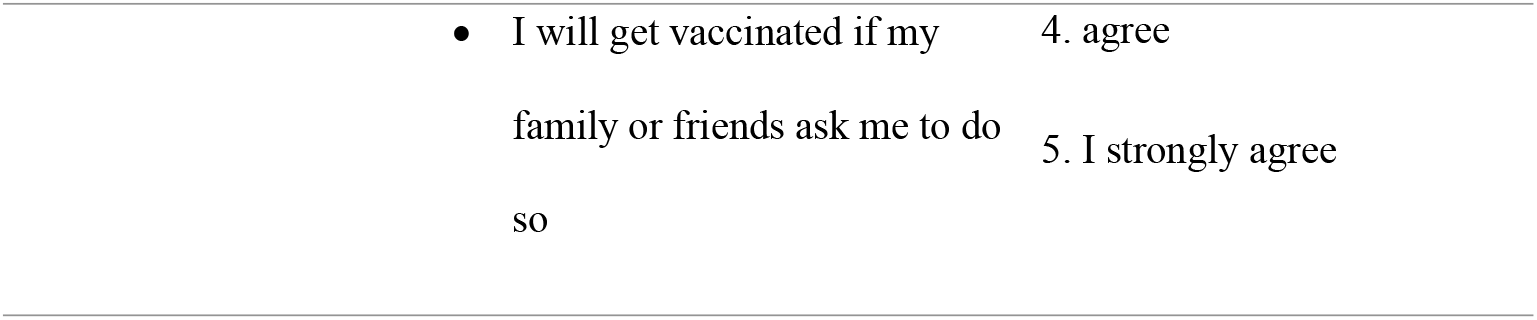

### 2.7 Sample size

The minimum sample size was calculated according to Schwartz’s formula [20].

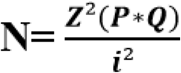

#### 2.7.1 Healthcare workers

We assumed that 50% of health workers are in favour of vaccination. With a desired accuracy of 5%, the minimum expected sample size is 384, and with 10% non-response rates, this size has been increased to 422 per city. A minimum of 1,688 women was required.

#### 2.7.2 General population

We assumed 20% vaccination coverage. With a desired accuracy of 5%, the minimum sample size was 245, and with 10% non-response rates, this size was increased to 270 per city. A minimum of 1080 women was required.

### 2.8 Data collection

The interviewers used Android phones to administer the questionnaire to participants at their workplace or by appointment at the nearest or most convenient location. The data was recorded via an Android application (ODK) downloaded and connected to the ONA server (https://ona.io/home/).

### 2.9 Ethical considerations

The study was approved by the Institutional Review Committee of Kofi Annan University of Guinea (020/UKAG/P9/2021). Health centre officials also consented before starting the investigation, and we collected data anonymously after obtaining informed consent from all participants. All methods were carried out under Guinean directives and regulations.

### 2.10 Statistical analysis

We transformed some variables before data analysis: perception/fear, attitudes and beliefs, subjective norms, ability and intention to receive the COVID-19 vaccine. To do this, we have made a classification according to the average of the scores of the scales.

- **Perception/fear:** we assumed participants with a score above or equal to the average were to have a positive perception. Otherwise, the perception is negative.
- **Attitude and belief:** divided into two parts:
  - Elements related to negative attitude: when the score is below average, the attitude is less negative; if necessary, the attitude is more pessimistic.
  - Elements related to the positive attitude: when the score is below average, the attitude is less positive; if necessary, the attitude is more positive.
- **Subjective norms:** when the score was below average, the standards were considered favourable; if so, the standards were unfavourable.
- **Ability to receive the COVID-19 vaccine:** When the score was below average, participants were considered incapable; if so, they were capable.
- **Intention to receive the COVID-19** vaccine: When the score is below average, participants have less intention to be vaccinated if necessary; they have a better vaccination intention.

We categorised variables and presented results as numbers and proportions. For HCWs and the GP, we tested the association between the dependent variable and each independent variable using Pearson’s Chi-squared and the Wilcoxon test.

We performed multivariate logistic regression to identify facilitators and barriers to participants’ vaccination acceptance. Indeed, we used a simple step-by-step procedure (without the possibility for a variable excluded at a previous step to be included later in the model) starting from the model containing all the independent variables and respecting the minimisation criterion of the Akaïke Information Criteria (AIC). We used the Hosmer-Lemeshow test to determine the fit quality of our regression model and tested Two-to-two interactions between independent variables. The adjusted odds ratio (ORa) with its 95% confidence interval (95% CI) measured the association between predictors and participants’ vaccination status.

We used variables from the final multivariate logistic regression model in the classification and regression tree (CART) to have a predictor profile of participants’ acceptance of a COVID-19 vaccine.

We performed analyses using R software version 4.1.2 and Stata 15 and considered statistical tests at risk α=5%.

## 3 Results

### 3.1 Analyse descriptive

We included 2,208 women for the HCWs and 1,121 for the GP (figure 1). Table 1 shows the sociodemographic characteristics of the HCWs and GP. We found that 54% of the HCWs and 45% of the GP knew about vaccination. Among the HCWs, 58% had researched COVID-19 compared to 47% of participants in GP. Most respondents negatively perceived the vaccine, with 53% for HCWs and 54% for GP, respectively. Attitudes were generally less negative, with 53% for the HCWs and 55% for the GP. Regarding positive attitudes, 71% of the HCWs had less positive attitudes, against 86% of the GP who had more positive attitudes.

**Table 1:**
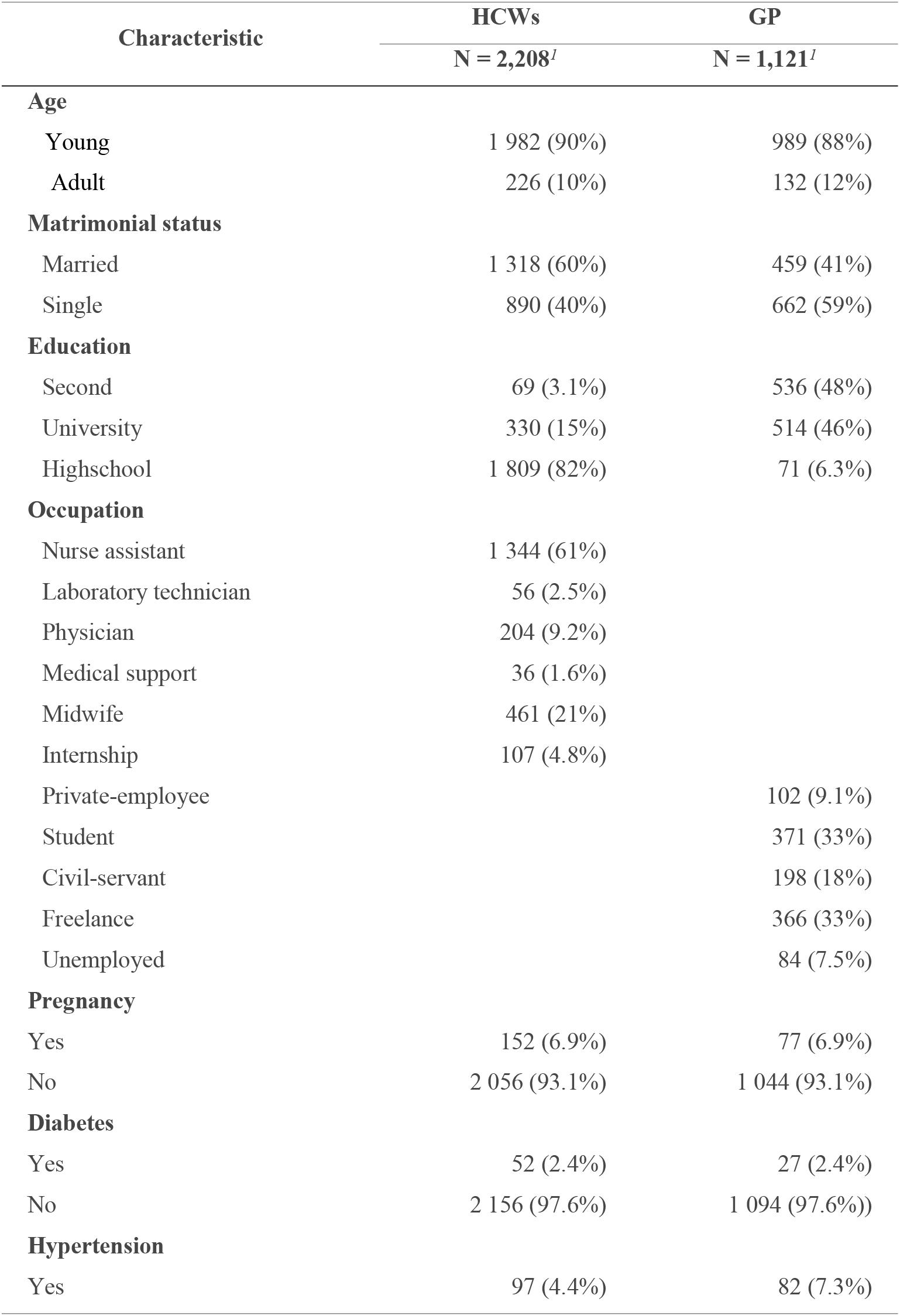
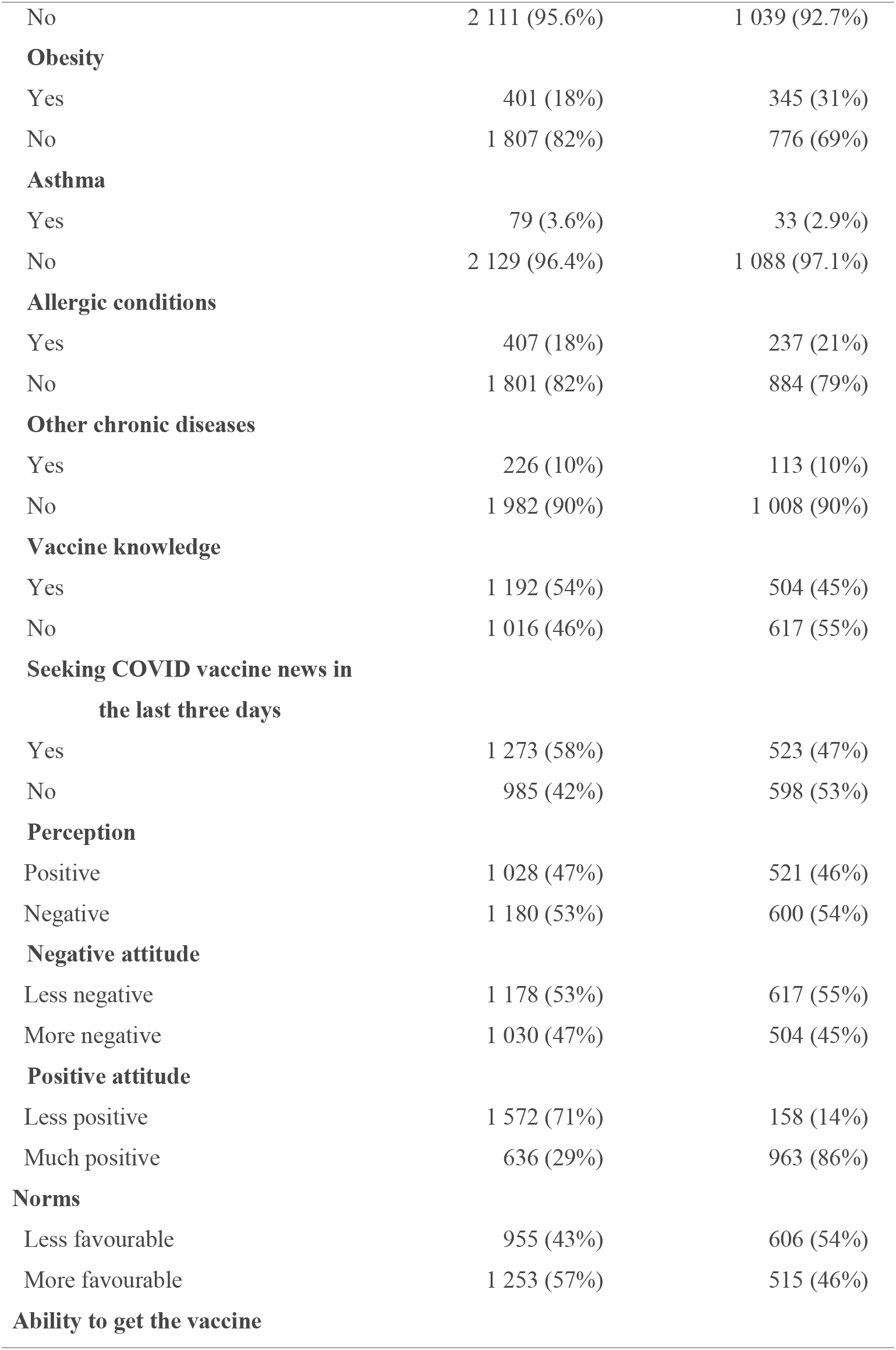
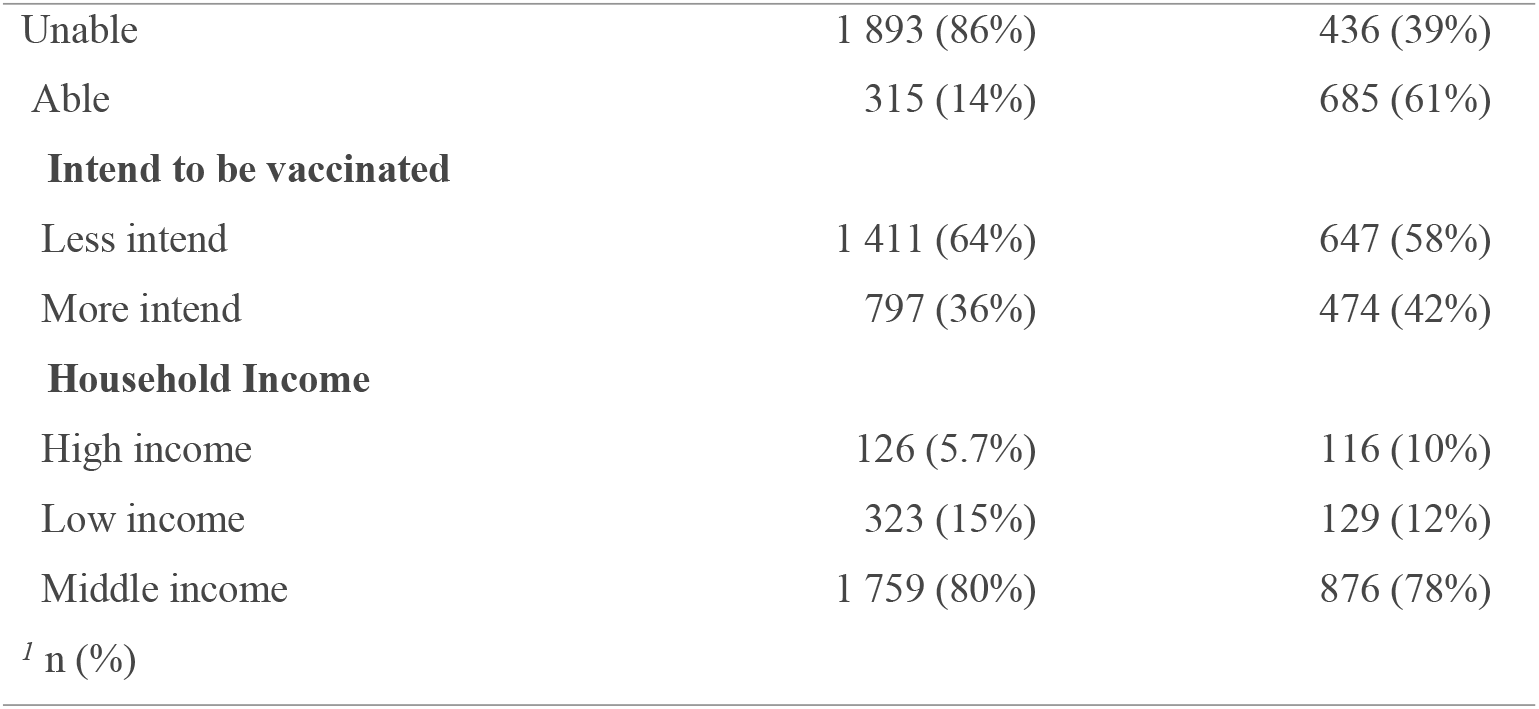
Sociodemographic characteristics of HCWs and GP participants. Guinea.2021

| Characteristic | HCWs | GP |
| --- | --- | --- |
|  | N = 2,208<sup>/</sup> | N = 1,121<sup>/</sup> |
| **Age** |  |  |
| Young | 1 982 (90%) | 989 (88%) |
| Adult | 226 (10%) | 132 (12%) |
| **Matrimonial status** |  |  |
| Married | 1 318 (60%) | 459 (41%) |
| Single | 890 (40%) | 662 (59%) |
| **Education** |  |  |
| Second | 69 (3.1%) | 536 (48%) |
| University | 330 (15%) | 514 (46%) |
| Highschool | 1 809 (82%) | 71 (6.3%) |
| **Occupation** |  |  |
| Nurse assistant | 1 344 (61%) |  |
| Laboratory technician | 56 (2.5%) |  |
| Physician | 204 (9.2%) |  |
| Medical support | 36 (1.6%) |  |
| Midwife | 461 (21%) |  |
| Internship | 107 (4.8%) |  |
| Private-employee |  | 102 (9.1%) |
| Student |  | 371 (33%) |
| Civil-servant |  | 198 (18%) |
| Freelance |  | 366 (33%) |
| Unemployed |  | 84 (7.5%) |
| **Pregnancy** |  |  |
| Yes | 152 (6.9%) | 77 (6.9%) |
| No | 2 056 (93.1%) | 1 044 (93.1%) |
| **Diabetes** |  |  |
| Yes | 52 (2.4%) | 27 (2.4%) |
| No | 2 156 (97.6%) | 1 094 (97.6%) |
| **Hypertension** |  |  |
| Yes | 97 (4.4%) | 82 (7.3%) |
| No | 2 111 (95.6%) | 1 039 (92.7%) |
| **Obesity** |  |  |
| Yes | 401 (18%) | 345 (31%) |
| No | 1 807 (82%) | 776 (69%) |
| **Asthma** |  |  |
| Yes | 79 (3.6%) | 33 (2.9%) |
| No | 2 129 (96.4%) | 1 088 (97.1%) |
| **Allergic conditions** |  |  |
| Yes | 407 (18%) | 237 (21%) |
| No | 1 801 (82%) | 884 (79%) |
| **Other chronic diseases** |  |  |
| Yes | 226 (10%) | 113 (10%) |
| No | 1 982 (90%) | 1 008 (90%) |
| **Vaccine knowledge** |  |  |
| Yes | 1 192 (54%) | 504 (45%) |
| No | 1 016 (46%) | 617 (55%) |
| **Seeking COVID vaccine news in**<br>**the last three days** |  |  |
| Yes | 1 273 (58%) | 523 (47%) |
| No | 985 (42%) | 598 (53%) |
| **Perception** |  |  |
| Positive | 1 028 (47%) | 521 (46%) |
| Negative | 1 180 (53%) | 600 (54%) |
| **Negative attitude** |  |  |
| Less negative | 1 178 (53%) | 617 (55%) |
| More negative | 1 030 (47%) | 504 (45%) |
| **Positive attitude** |  |  |
| Less positive | 1 572 (71%) | 158 (14%) |
| Much positive | 636 (29%) | 963 (86%) |
| **Norms** |  |  |
| Less favourable | 955 (43%) | 606 (54%) |
| More favourable | 1 253 (57%) | 515 (46%) |
| **Ability to get the vaccine** |  |  |
| Unable | 1 893 (86%) | 436 (39%) |
| Able | 315 (14%) | 685 (61%) |
| **Intend to be vaccinated** |  |  |
| Less intend | 1 411 (64%) | 647 (58%) |
| More intend | 797 (36%) | 474 (42%) |
| **Household Income** |  |  |
| High income | 126 (5.7%) | 116 (10%) |
| Low income | 323 (15%) | 129 (12%) |
| Middle income | 1 759 (80%) | 876 (78%) |
| <sup>1</sup> n (%) |  |  |

**Figure 1:**
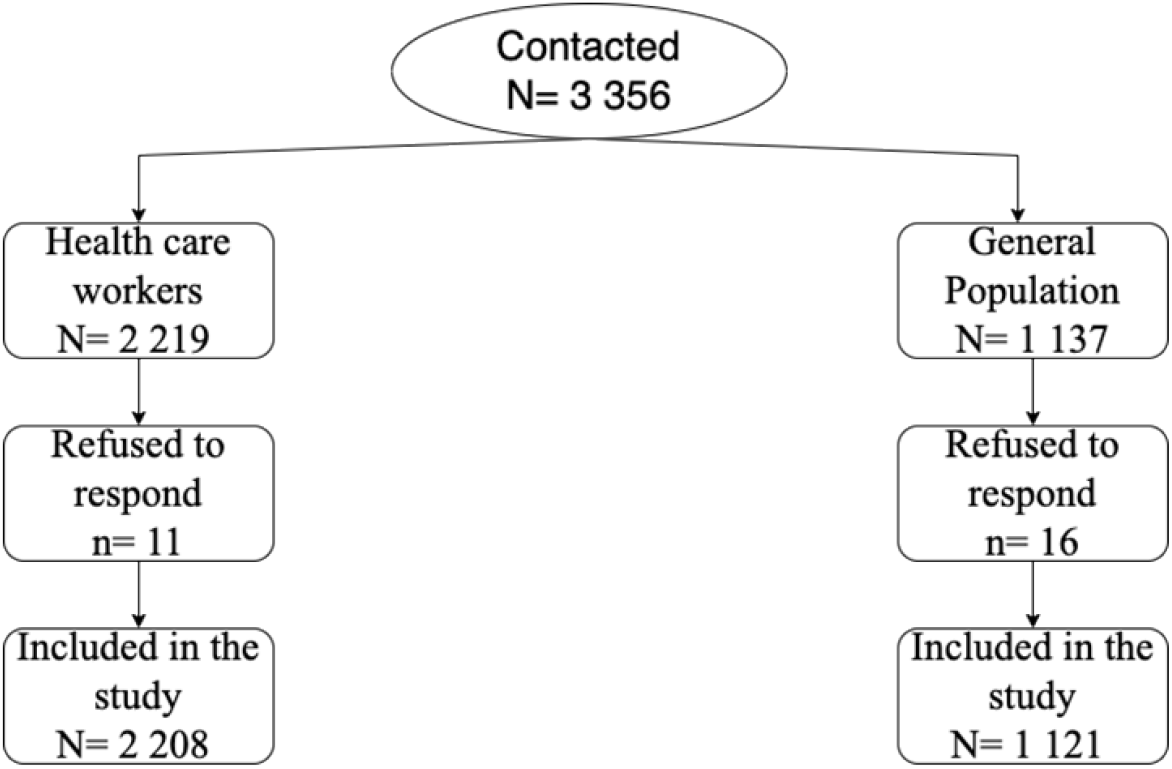
Inclusion Flow Diagram

Most participants had low vaccination intention, 64% for HCWs and 58% for GP. Incomes mainly were intermediate, 80% for the HCWs and 78% for the GP. Of the PS, 86% could not receive the vaccine. However, 61% of the PG had the capacity. Subjective norms were favourable in 57% of cases for the HCWs and less favourable in 54% of cases for the GP.

### 3.2 Sources of information

Figure 2 shows that the primary source of information for the HCWs participants was social networks, with 935 cases (42.35%).

**Figure 2:**
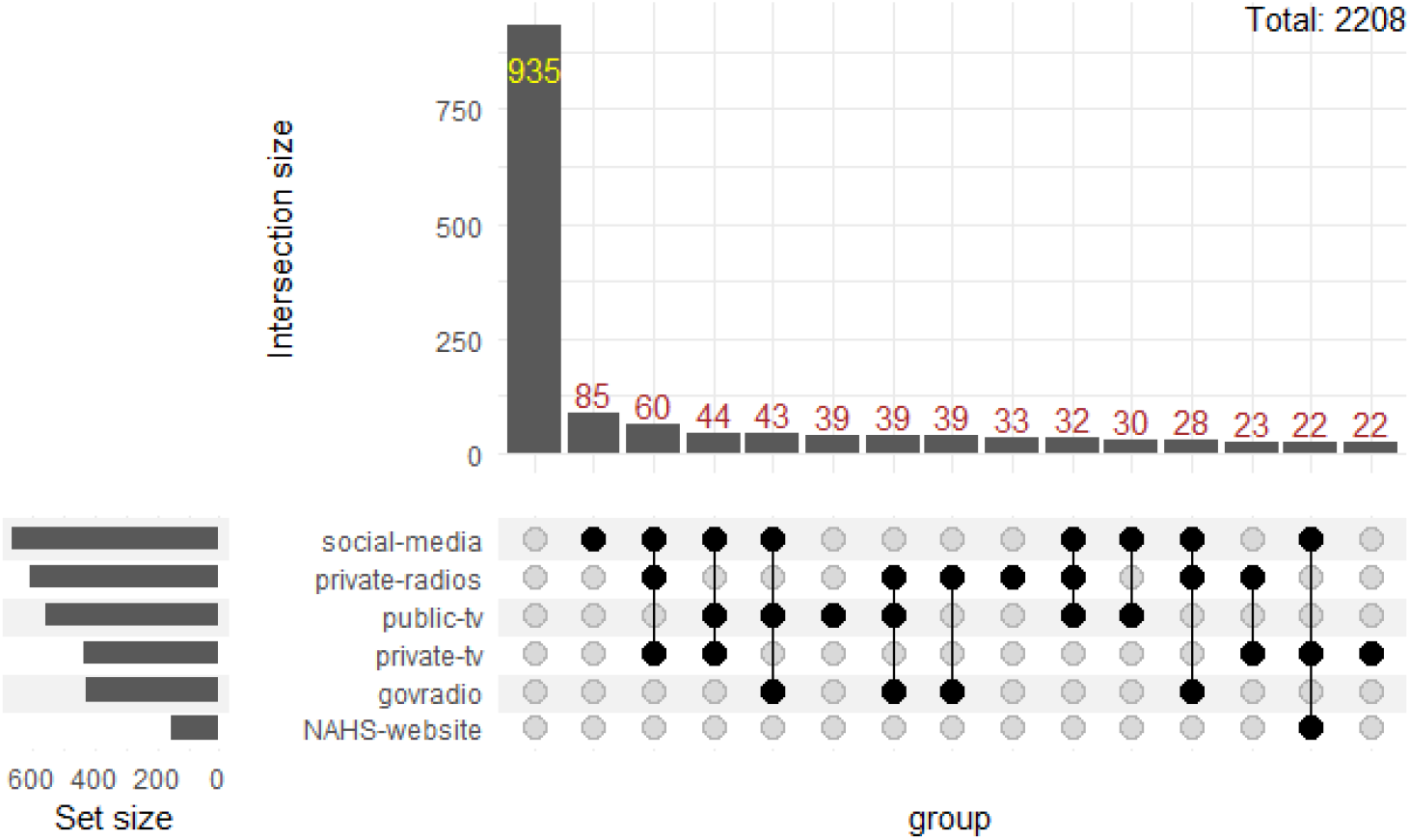
HCWs COVID-19 information sources. Guinea.2021

Figure 3 shows that GP participants had social networks as their primary source of information, with 598 cases (53.35%).

**Figure 3:**
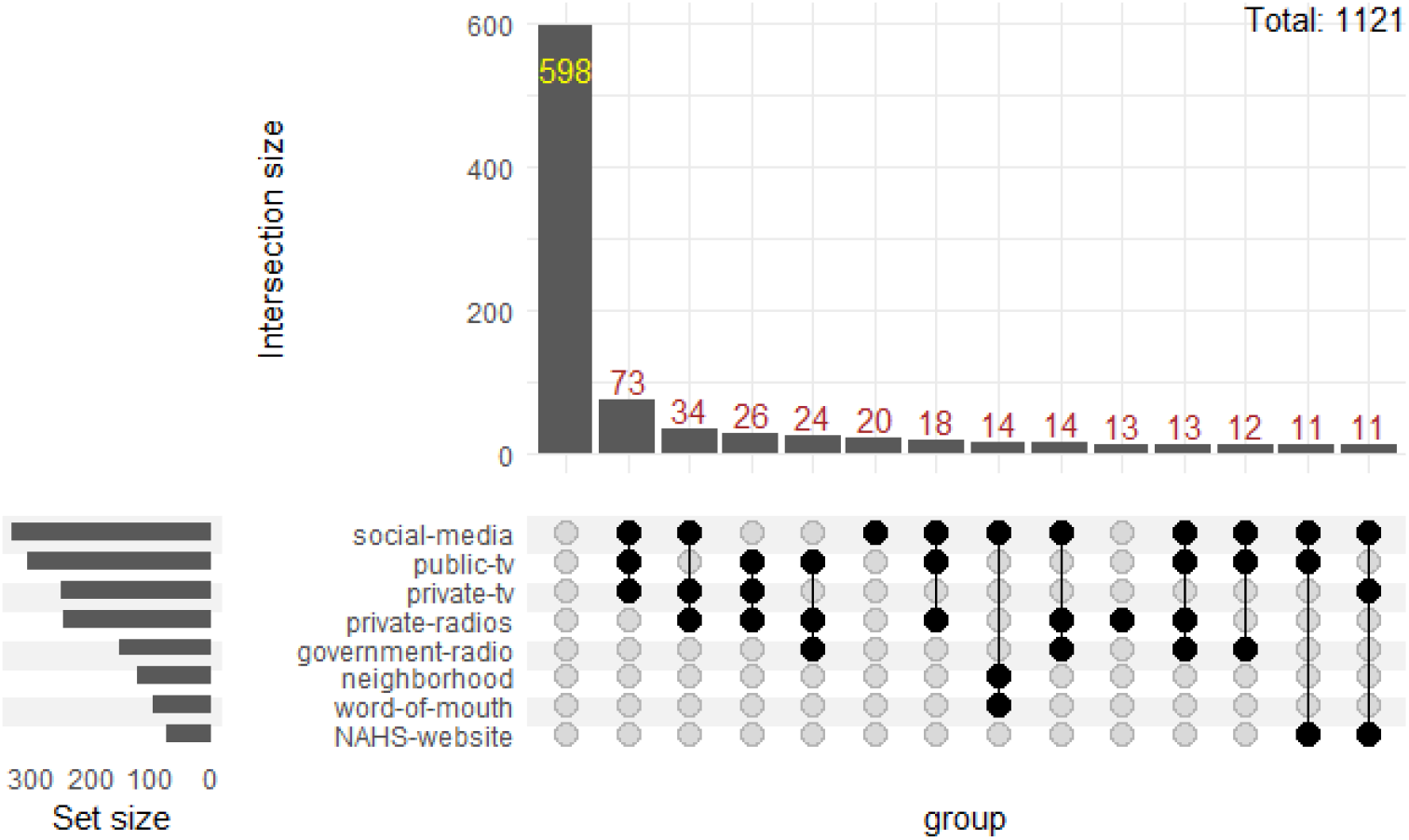
GP COVID-19 information sources. Guinea.2021

### 3.3 Vaccination rate (HCWs)

La figure 4 shows that most HCWs respondents were vaccinated (63%).

**Figure 4:**
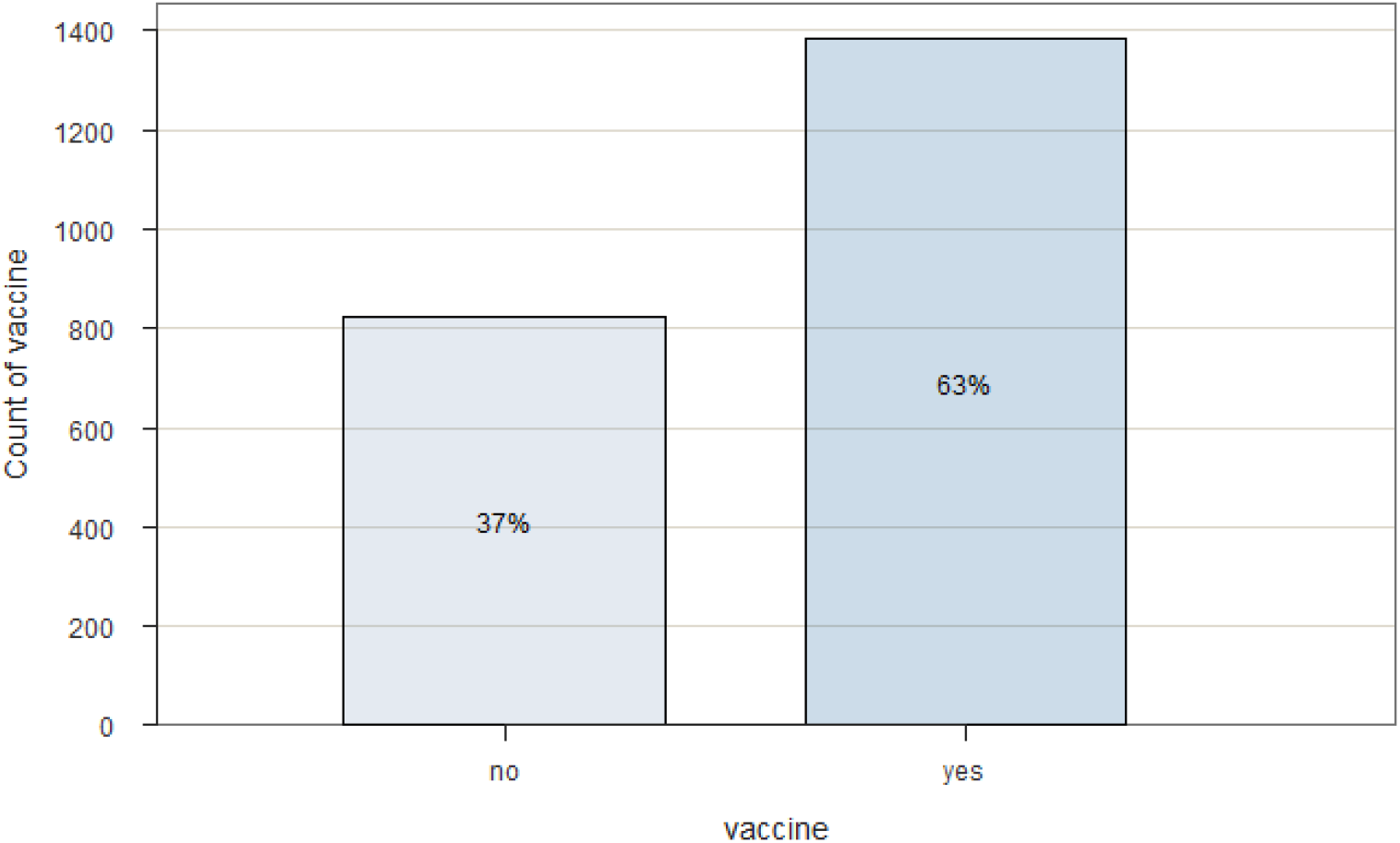
Proportion of HCWs participants vaccinated against COVID-19. From Mar 22 to Aug 25 2021. Guinea. N=2,208.

### General population

Figure 5 shows that only 28% of GP participants were vaccinated.

**Figure 5:**
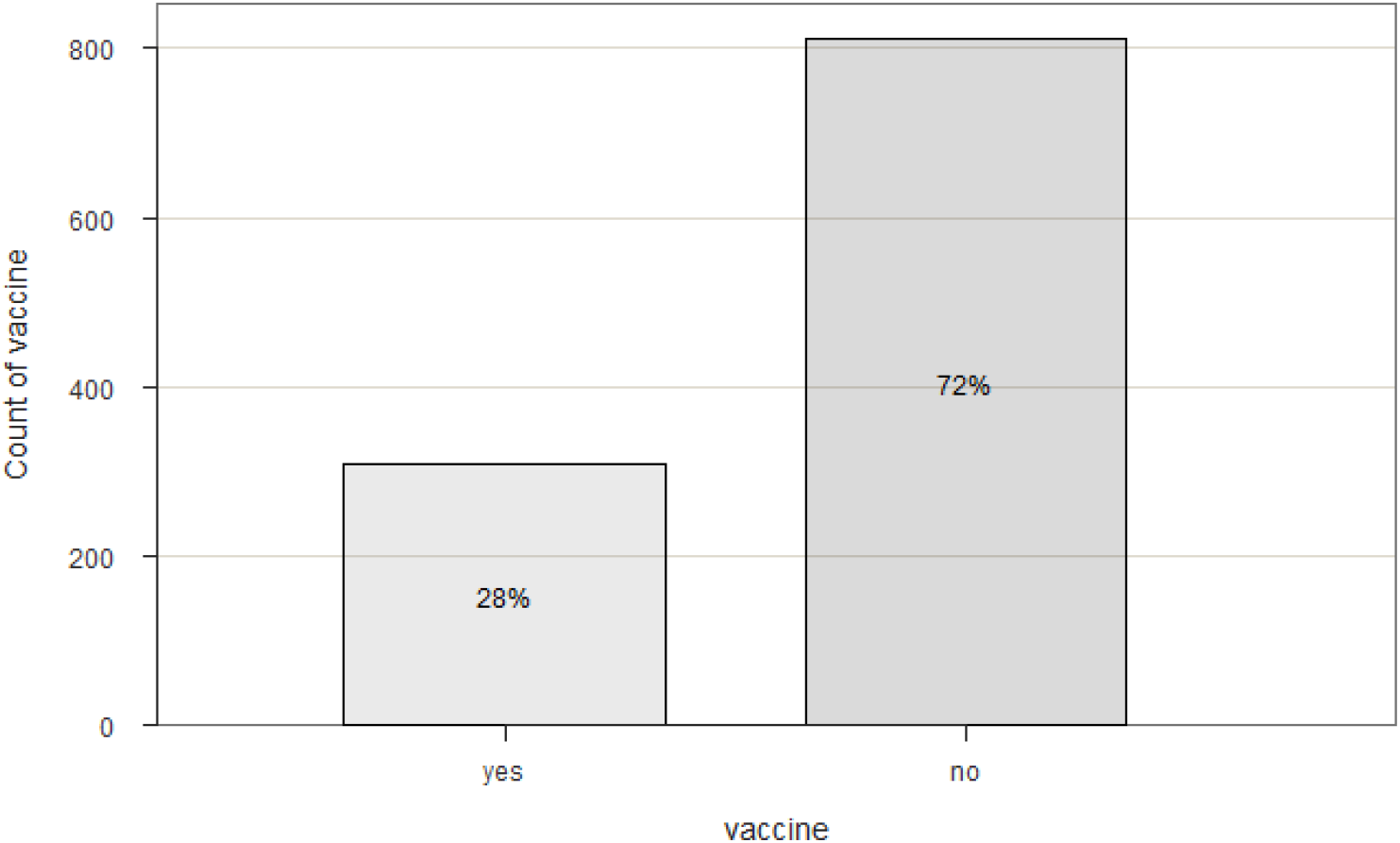
Proportion of GP participants vaccinated against COVID-19. From Mar 22 to Aug 25 2021. Guinea. N=1,121.

### 3.4 Univariate analysis

Table 2 shows the factors significantly associated with COVID-19 vaccination.

**Table 2:**
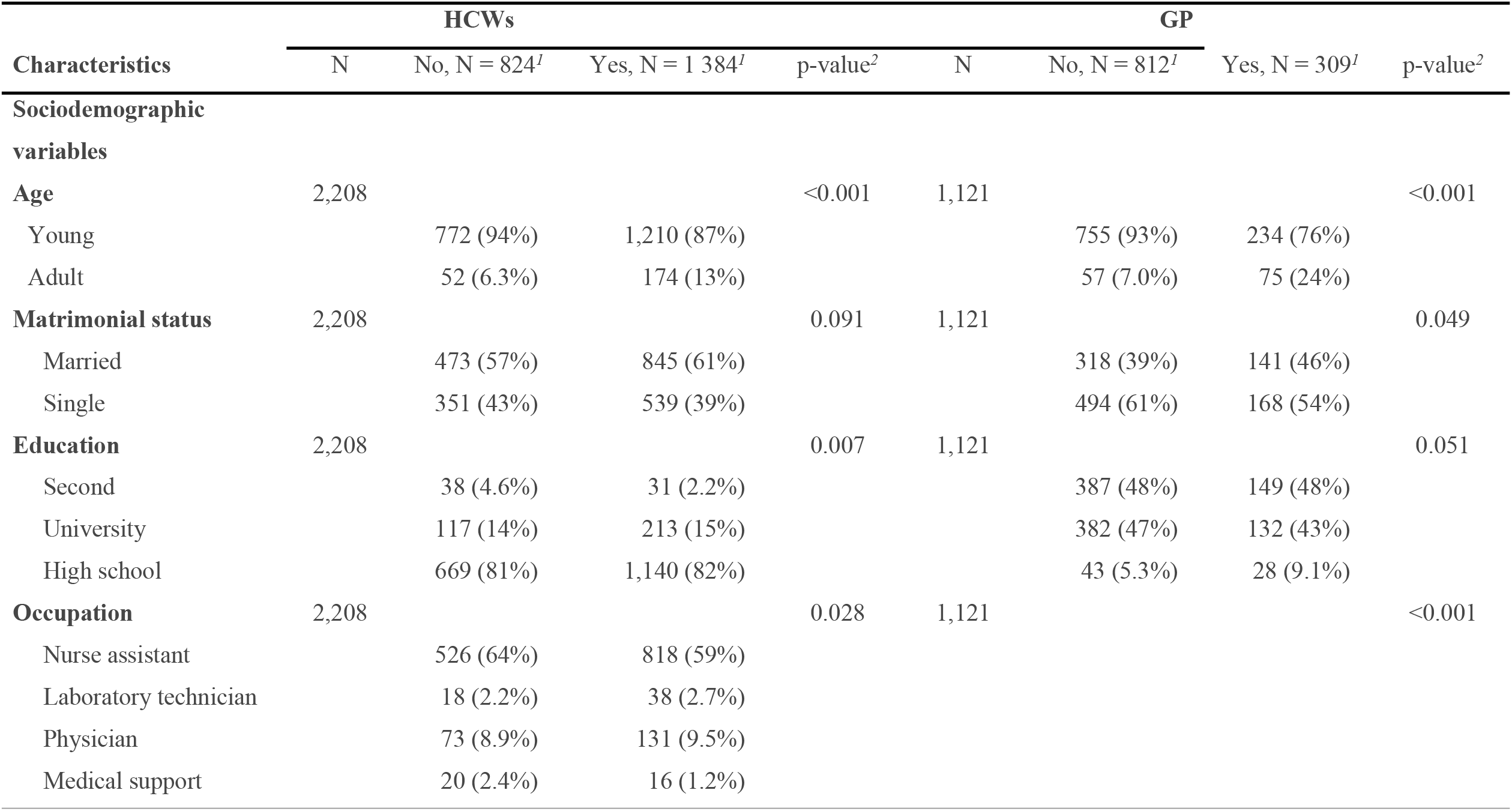
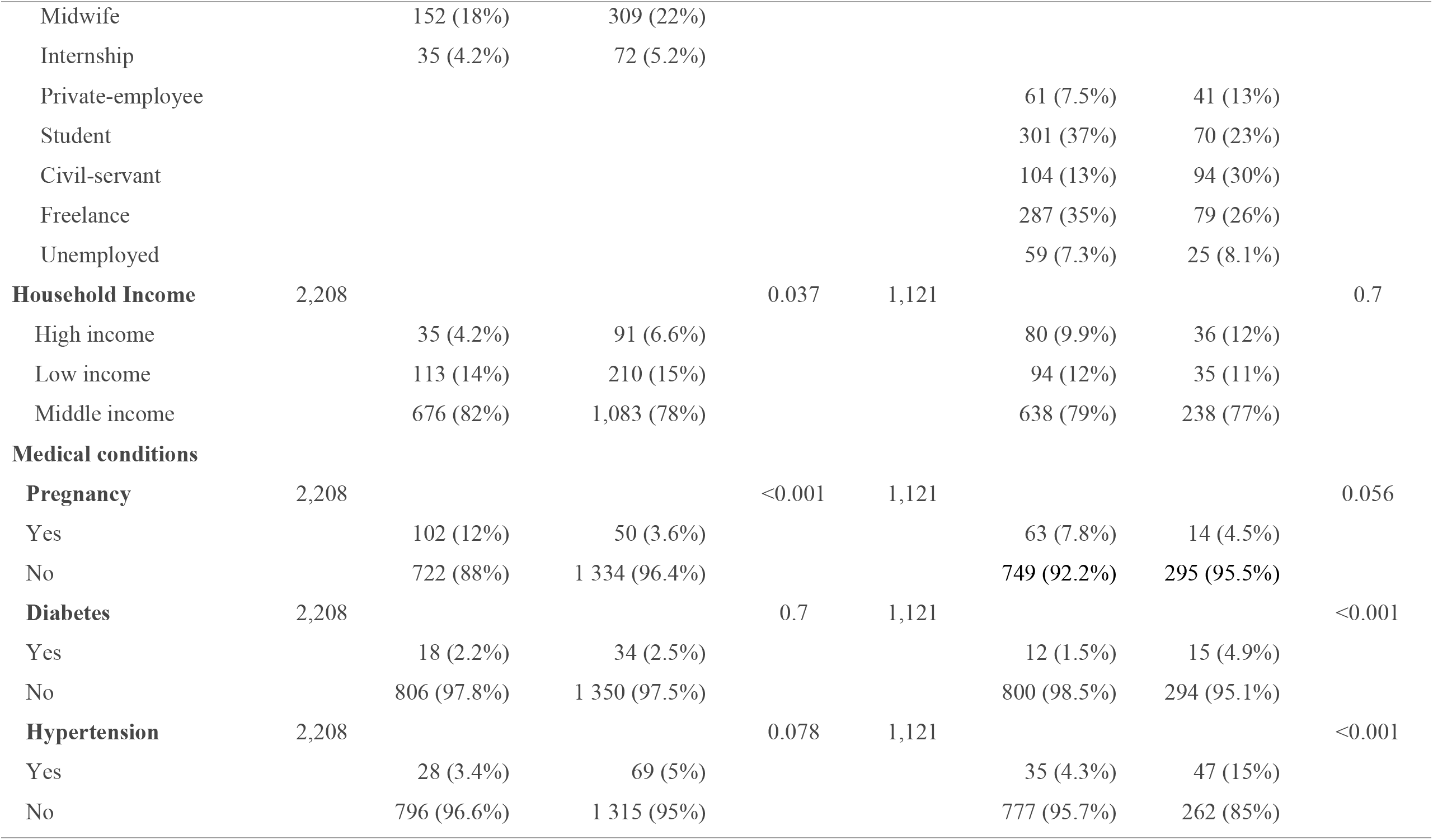
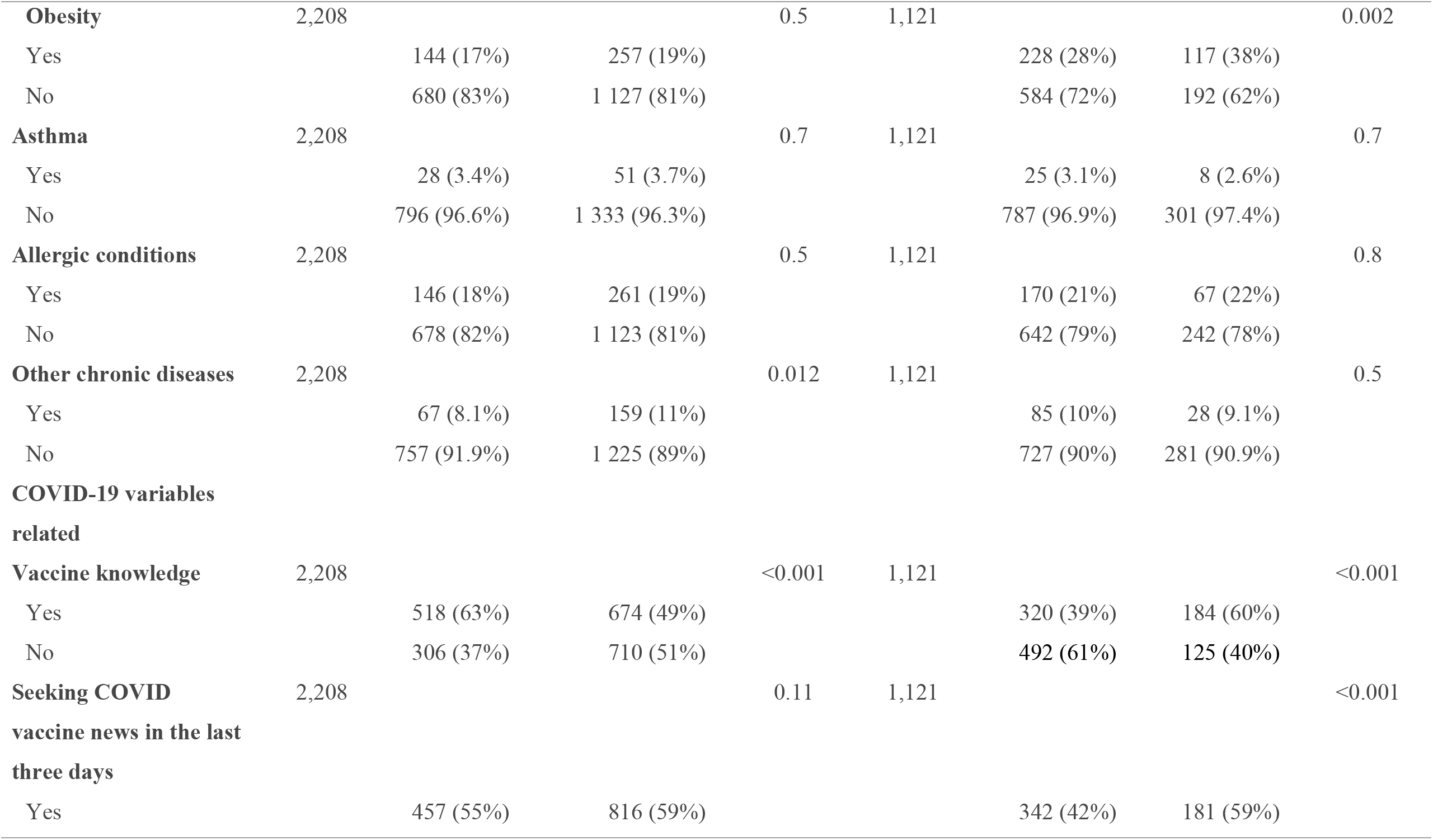
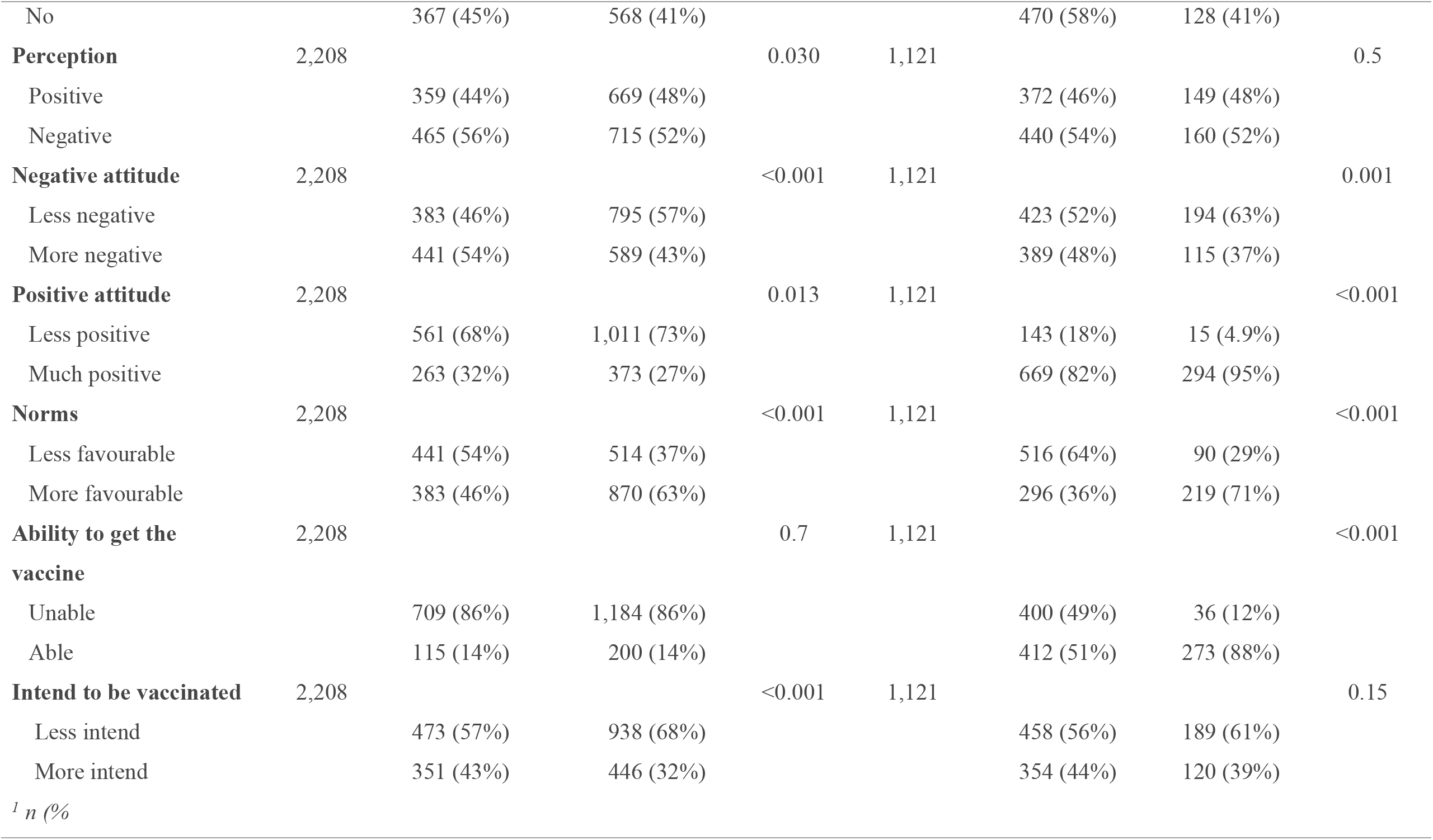
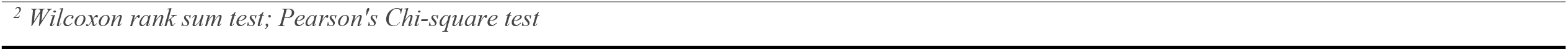
Univariate analysis of acceptance of COVID-19 vaccination by **HCWs a**nd GP. From Mar 22 to Aug 25 2021. Guinea.

| Characteristics | HCWs |  |  |  | GP |  |  |  |
| --- | --- | --- | --- | --- | --- | --- | --- | --- |
|  | N | No, N = 824<sup>I</sup> | Yes, N = 1 384<sup>I</sup> | p-value<sup>2</sup> | N | No, N = 812<sup>I</sup> | Yes, N = 309<sup>I</sup> | p-value<sup>2</sup> |
| **Sociodemographic variables** |  |  |  |  |  |  |  |  |
| **Age** | 2,208 |  |  | <0.001 | 1,121 |  |  | <0.001 |
| Young |  | 772 (94%) | 1,210 (87%) |  |  | 755 (93%) | 234 (76%) |  |
| Adult |  | 52 (6.3%) | 174 (13%) |  |  | 57 (7.0%) | 75 (24%) |  |
| **Matrimonial status** | 2,208 |  |  | 0.091 | 1,121 |  |  | 0.049 |
| Married |  | 473 (57%) | 845 (61%) |  |  | 318 (39%) | 141 (46%) |  |
| Single |  | 351 (43%) | 539 (39%) |  |  | 494 (61%) | 168 (54%) |  |
| **Education** | 2,208 |  |  | 0.007 | 1,121 |  |  | 0.051 |
| Second |  | 38 (4.6%) | 31 (2.2%) |  |  | 387 (48%) | 149 (48%) |  |
| University |  | 117 (14%) | 213 (15%) |  |  | 382 (47%) | 132 (43%) |  |
| High school |  | 669 (81%) | 1,140 (82%) |  |  | 43 (5.3%) | 28 (9.1%) |  |
| **Occupation** | 2,208 |  |  | 0.028 | 1,121 |  |  | <0.001 |
| Nurse assistant |  | 526 (64%) | 818 (59%) |  |  |  |  |  |
| Laboratory technician |  | 18 (2.2%) | 38 (2.7%) |  |  |  |  |  |
| Physician |  | 73 (8.9%) | 131 (9.5%) |  |  |  |  |  |
| Medical support |  | 20 (2.4%) | 16 (1.2%) |  |  |  |  |  |
| Midwife |  | 152 (18%) | 309 (22%) |  |  |  |  |  |
| Internship |  | 35 (4.2%) | 72 (5.2%) |  |  |  |  |  |
| Private-employee |  |  |  |  |  | 61 (7.5%) | 41 (13%) |  |
| Student |  |  |  |  |  | 301 (37%) | 70 (23%) |  |
| Civil-servant |  |  |  |  |  | 104 (13%) | 94 (30%) |  |
| Freelance |  |  |  |  |  | 287 (35%) | 79 (26%) |  |
| Unemployed |  |  |  |  |  | 59 (7.3%) | 25 (8.1%) |  |
| **Household Income** | 2,208 |  |  | 0.037 | 1,121 |  |  | 0.7 |
| High income |  | 35 (4.2%) | 91 (6.6%) |  |  | 80 (9.9%) | 36 (12%) |  |
| Low income |  | 113 (14%) | 210 (15%) |  |  | 94 (12%) | 35 (11%) |  |
| Middle income |  | 676 (82%) | 1,083 (78%) |  |  | 638 (79%) | 238 (77%) |  |
| **Medical conditions** |  |  |  |  |  |  |  |  |
| **Pregnancy** | 2,208 |  |  | <0.001 | 1,121 |  |  | 0.056 |
| Yes |  | 102 (12%) | 50 (3.6%) |  |  | 63 (7.8%) | 14 (4.5%) |  |
| No |  | 722 (88%) | 1 334 (96.4%) |  |  | 749 (92.2%) | 295 (95.5%) |  |
| **Diabetes** | 2,208 |  |  | 0.7 | 1,121 |  |  | <0.001 |
| Yes |  | 18 (2.2%) | 34 (2.5%) |  |  | 12 (1.5%) | 15 (4.9%) |  |
| No |  | 806 (97.8%) | 1 350 (97.5%) |  |  | 800 (98.5%) | 294 (95.1%) |  |
| **Hypertension** | 2,208 |  |  | 0.078 | 1,121 |  |  | <0.001 |
| Yes |  | 28 (3.4%) | 69 (5%) |  |  | 35 (4.3%) | 47 (15%) |  |
| No |  | 796 (96.6%) | 1 315 (95%) |  |  | 777 (95.7%) | 262 (85%) |  |
| **Obesity** | 2,208 |  |  | 0.5 | 1,121 |  |  | 0.002 |
| Yes |  | 144 (17%) | 257 (19%) |  |  | 228 (28%) | 117 (38%) |  |
| No |  | 680 (83%) | 1 127 (81%) |  |  | 584 (72%) | 192 (62%) |  |
| **Asthma** | 2,208 |  |  | 0.7 | 1,121 |  |  | 0.7 |
| Yes |  | 28 (3.4%) | 51 (3.7%) |  |  | 25 (3.1%) | 8 (2.6%) |  |
| No |  | 796 (96.6%) | 1 333 (96.3%) |  |  | 787 (96.9%) | 301 (97.4%) |  |
| **Allergic conditions** | 2,208 |  |  | 0.5 | 1,121 |  |  | 0.8 |
| Yes |  | 146 (18%) | 261 (19%) |  |  | 170 (21%) | 67 (22%) |  |
| No |  | 678 (82%) | 1 123 (81%) |  |  | 642 (79%) | 242 (78%) |  |
| **Other chronic diseases** | 2,208 |  |  | 0.012 | 1,121 |  |  | 0.5 |
| Yes |  | 67 (8.1%) | 159 (11%) |  |  | 85 (10%) | 28 (9.1%) |  |
| No |  | 757 (91.9%) | 1 225 (89%) |  |  | 727 (90%) | 281 (90.9%) |  |
| **COVID-19 variables related** |  |  |  |  |  |  |  |  |
| **Vaccine knowledge** | 2,208 |  |  | <0.001 | 1,121 |  |  | <0.001 |
| Yes |  | 518 (63%) | 674 (49%) |  |  | 320 (39%) | 184 (60%) |  |
| No |  | 306 (37%) | 710 (51%) |  |  | 492 (61%) | 125 (40%) |  |
| **Seeking COVID vaccine news in the last three days** | 2,208 |  |  | 0.11 | 1,121 |  |  | <0.001 |
| Yes |  | 457 (55%) | 816 (59%) |  |  | 342 (42%) | 181 (59%) |  |
| No |  | 367 (45%) | 568 (41%) |  |  | 470 (58%) | 128 (41%) |  |
| **Perception** | 2,208 |  |  | 0.030 | 1,121 |  |  | 0.5 |
| Positive |  | 359 (44%) | 669 (48%) |  |  | 372 (46%) | 149 (48%) |  |
| Negative |  | 465 (56%) | 715 (52%) |  |  | 440 (54%) | 160 (52%) |  |
| **Negative attitude** | 2,208 |  |  | <0.001 | 1,121 |  |  | 0.001 |
| Less negative |  | 383 (46%) | 795 (57%) |  |  | 423 (52%) | 194 (63%) |  |
| More negative |  | 441 (54%) | 589 (43%) |  |  | 389 (48%) | 115 (37%) |  |
| **Positive attitude** | 2,208 |  |  | 0.013 | 1,121 |  |  | <0.001 |
| Less positive |  | 561 (68%) | 1,011 (73%) |  |  | 143 (18%) | 15 (4.9%) |  |
| Much positive |  | 263 (32%) | 373 (27%) |  |  | 669 (82%) | 294 (95%) |  |
| **Norms** | 2,208 |  |  | <0.001 | 1,121 |  |  | <0.001 |
| Less favourable |  | 441 (54%) | 514 (37%) |  |  | 516 (64%) | 90 (29%) |  |
| More favourable |  | 383 (46%) | 870 (63%) |  |  | 296 (36%) | 219 (71%) |  |
| **Ability to get the vaccine** | 2,208 |  |  | 0.7 | 1,121 |  |  | <0.001 |
| Unable |  | 709 (86%) | 1,184 (86%) |  |  | 400 (49%) | 36 (12%) |  |
| Able |  | 115 (14%) | 200 (14%) |  |  | 412 (51%) | 273 (88%) |  |
| **Intend to be vaccinated** | 2,208 |  |  | <0.001 | 1,121 |  |  | 0.15 |
| Less intend |  | 473 (57%) | 938 (68%) |  |  | 458 (56%) | 189 (61%) |  |
| More intend |  | 351 (43%) | 446 (32%) |  |  | 354 (44%) | 120 (39%) |  |
| <sup>1</sup> n (%) |  |  |  |  |  |  |  |  |

<sup>2</sup> *Wilcoxon rank sum test; Pearson's Chi-square test*

**HCWs:** factors associated with vaccination were age, education, occupation, pregnancy, immunisation knowledge, perception, negative attitudes, positive attitudes, subjective norms, intention to get vaccinated, and income.

**GP:** factors associated with vaccination were age, marital status, occupation, diabetes, hypertension, obesity, seeking information about COVID-19, knowledge about vaccination, subjective norms, negative attitudes, positive attitudes, and ability to receive the vaccine.

### 3.5 Multivariate analysis

Table 3 shows predictors of acceptance of vaccination by HCP and PG.

**Table 3:**
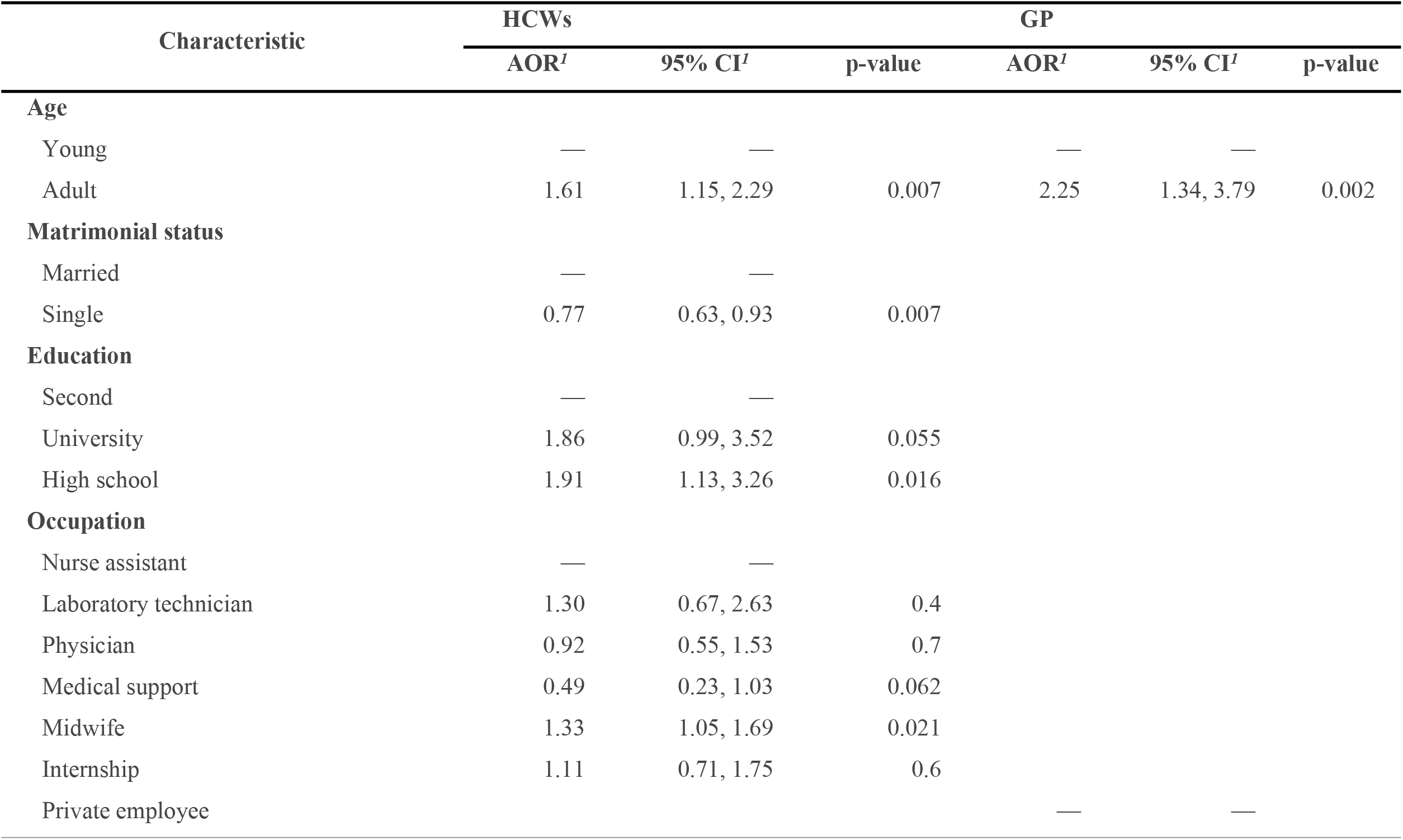
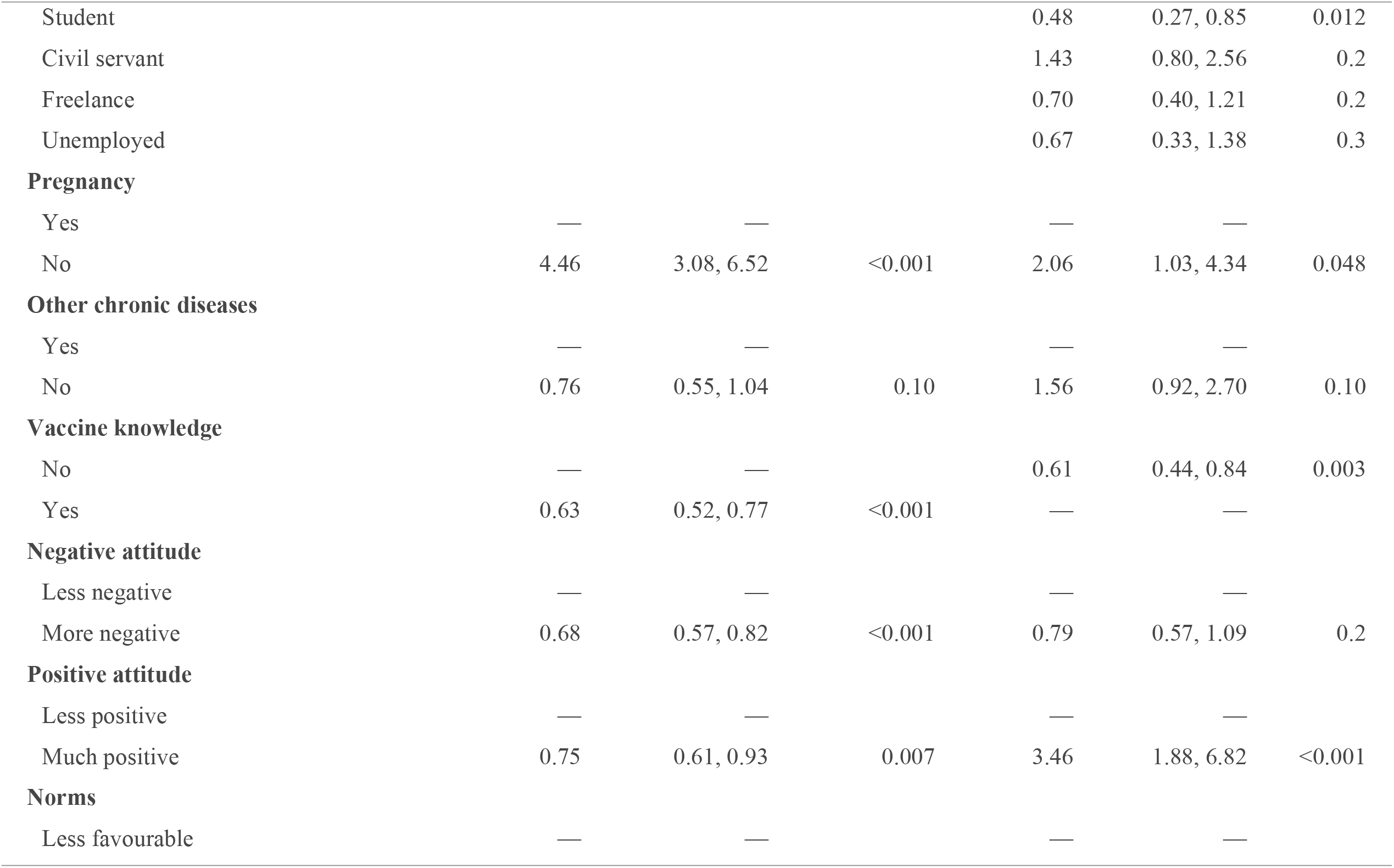
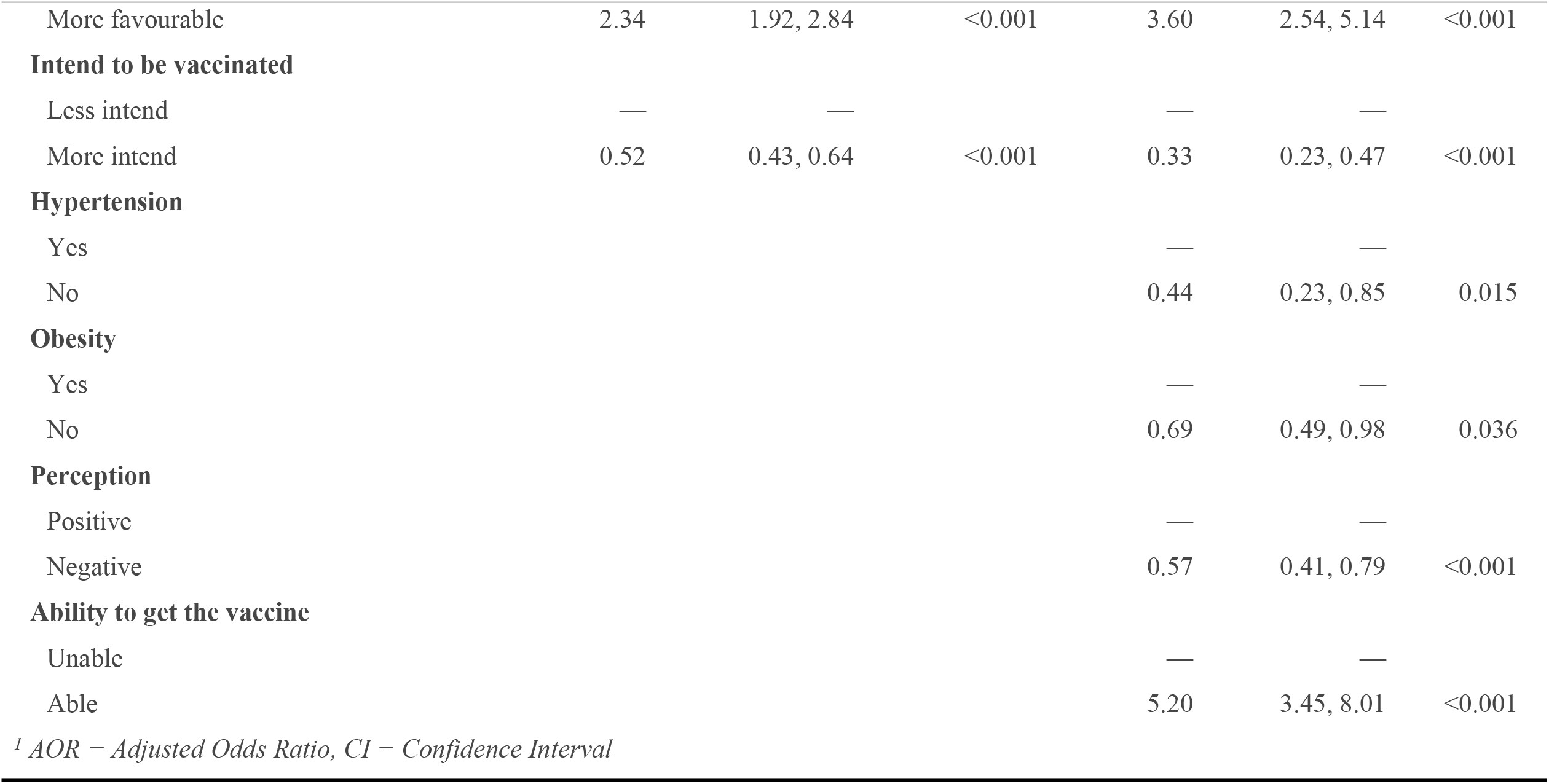
Multivariate analysis of acceptance of COVID-19 vaccination by HCP and PG. From Mar 22 to Aug 25 2021. Guinea

#### HCWs

Adults were 1.61 times more likely to accept vaccination than young AOR=1.61 95% [CI: 1.15, 2.29]. Single women were less likely to get vaccination than married women AOR =0.77 95% CI [0.63, 0.93]. Participants with a professional level of education were significantly 1.91 times more likely to accept vaccination than those in secondary AOR =1.91 95% CI [1.13, 3.26]. Midwives were 1.33 times more likely to get vaccination than nurse assistants AOR =1.33 95% CI [1.05, 1.69]. Vaccine acceptance was 4.46 times higher in non-pregnant women than in pregnant women AOR =4.46 95% CI [3.08, 6.52]. HCWs who knew about vaccination were less likely to accept the COVID-19 vaccine than those who did not know AOR =0.63 95% CI [0.52, 0.77]. Women with a more negative attitude were less likely to be vaccinated than those with a less negative attitude AOR =0.68 95% CI [0.57, 0.82]. Similarly, workers with more positive attitudes were less likely to be vaccinated than those with less positive attitudes AOR =0.75 CI95% [0.61, 0.93]. Respondents with favourable subjective norms were 2.34 times more likely to accept vaccination than those with less favourable subjective norms AOR =2.34 95% CI [1.92, 2.84]. Participants with a better vaccination intention were less likely to get the COVID-19 vaccine than those with a low ORA intention = 0.52 95% CI [0.43, 0.64].

#### GP

Adults were 2.25 times more likely to accept the COVID-19 vaccine than young ORA=2.25 95% CI [1.34, 3.79]. Female students were less likely to get the COVID-19 vaccine than private employees ORA=0.48 95% CI [0.27, 0.85]. Participants with a more positive attitude were 3.36 times more likely to be vaccinated than those with a less positive attitude ORA=3.4695% CI [1.88, 6.82]. Workers with a better vaccination intention were less likely to accept the COVID-19 vaccine than those with a low ORA line-of=0.33 95% CI [0.23, 0.47].

Non-pregnant women were 2.06 times more likely to accept vaccination than pregnant ORA=2.06 95% CI [1.03, 4.34]. Non-hypertensive participants were less likely to get the COVID-19 vaccine than those with hypertension ORA=0.44 95% CI [0.23, 0.85]. Similarly, non-obese women were less likely to accept vaccination than obese ORA=0.69 95% CI [0.49, 0.98]. Participants without good immunisation knowledge were less likely to get vaccination than those with good knowledge ORA=0.61 CI 95% [0.44, 0.84].

Women with a negative perception of the vaccine were less likely to accept vaccination than those with a positive ORA=0.57 95% CI [0.41, 0.79]. Participants with favourable subjective norms were 3.60 times more likely to get vaccination than those with less favourable subjective norms ORA=3.60 95% CI [2.54, 5.14]. Women who could receive the vaccine were 5.20 times more likely to accept COVID-19 vaccination than those who did not have the ORA capacity = 5.20 95% CI [3.45, 8.01].

#### Classification and regression tree (CART)

##### Healthcare workers

Figure 6 shows that pregnancy was the main factor distinguishing five (5) groups. The acceptance of vaccination was lower among pregnant women. Alternatively, it was better in non-pregnant women with favourable subjective norms and lower vaccination intention.

**Figure 6:**
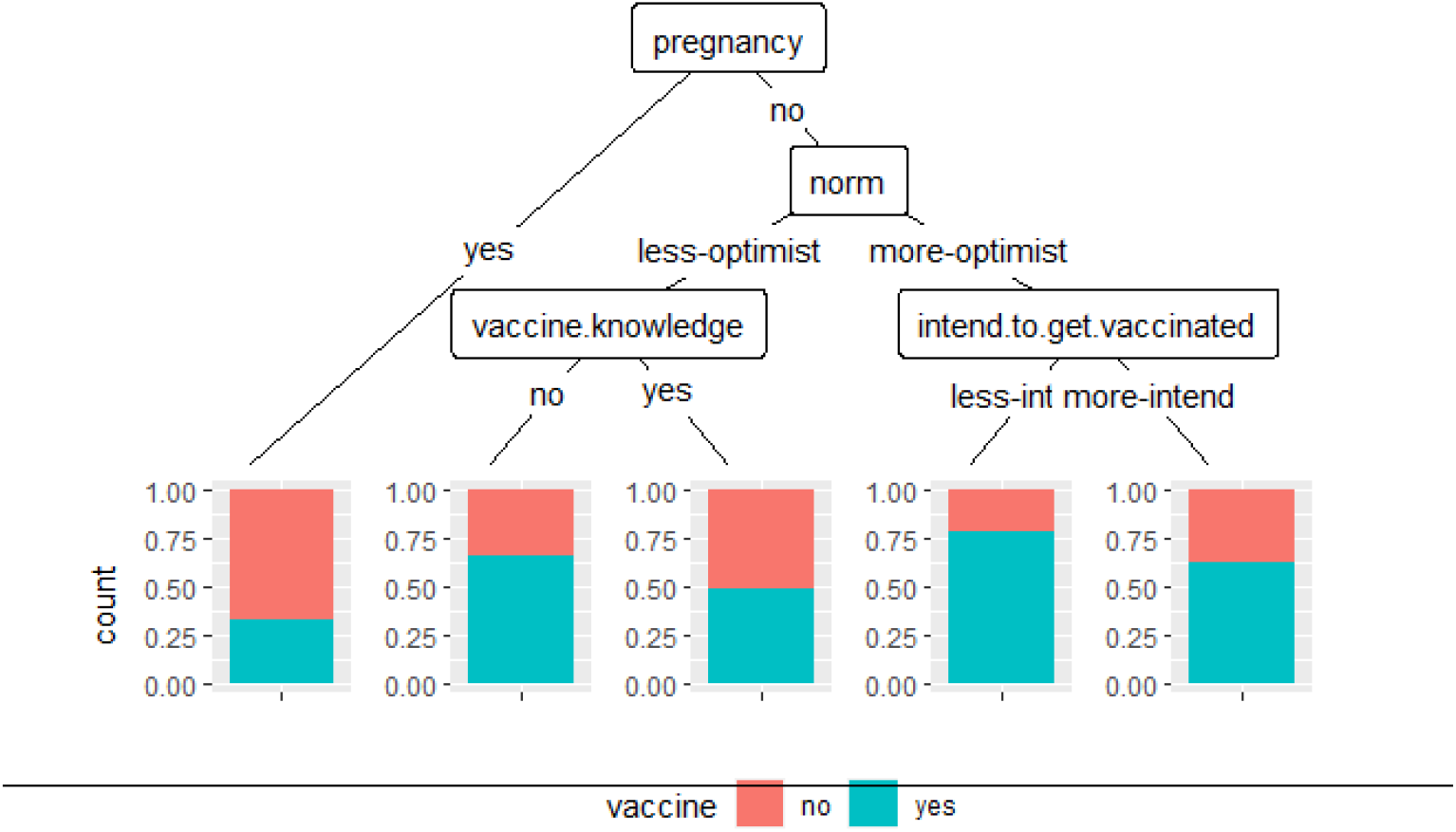
Predictor profiles of HCWs’ acceptance of COVID-19 vaccination with CART. From Mar 22 to Aug 25 2021. Guinea.

#### General population

Figure 7 shows that the ability to be vaccinated was the main factor in distinguishing four (4) groups. Vaccination acceptance was shallow among women who could not receive the vaccine and had less favourable subjective norms. However, approval was better among women who could receive the vaccine and were adults.

**Figure 7:**
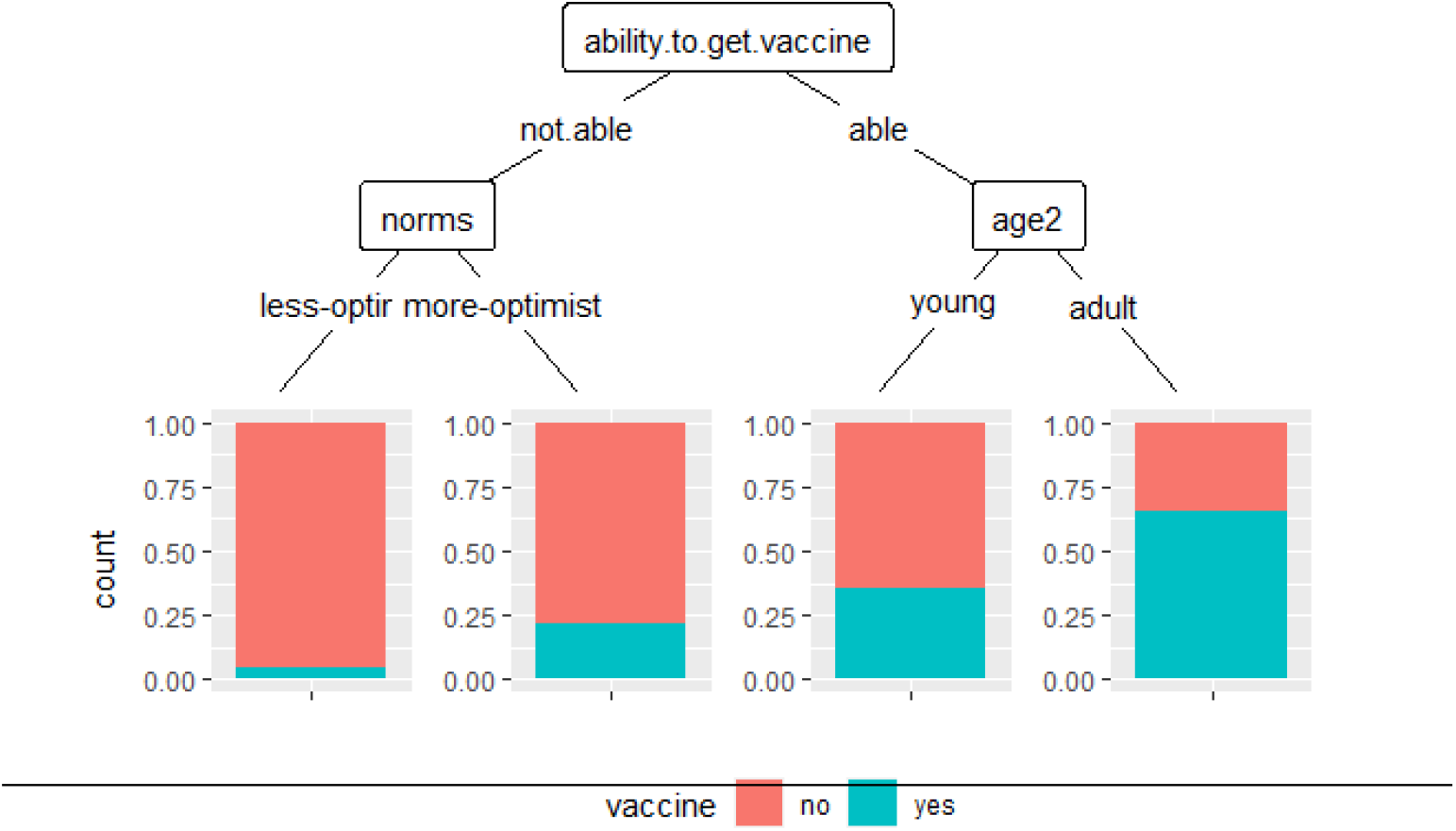
Predictor profiles of acceptance of COVID-19 vaccination by GP with CART. From Mar 22 to Aug 25 2021. Guinea.

## 4 Discussion

In this study, we sought to determine the acceptability and hesitation of the COVID-19 vaccine among women in Guinea and to identify the main associated predictors. Our results indicated that closer to 2/3 of healthcare workers were already vaccinated. Indeed, from the first vaccination phase, the national strategy, “Targeted Vaccination”, prioritised frontline health workers [21]. However, in our study, there were still just over a third of unvaccinated health workers. This situation would indicate hesitation or refusal of vaccination. The reluctance or refusal of vaccination among healthcare workers is believed to stem mainly from doubts about the safety and effectiveness of COVID-19 vaccines [22].

In the general population, less than one-third of our participants were vaccinated. Vaccinating at least 60% of the Guinean population with a safe and effective vaccine is insufficient by 2022 [23]. Furthermore, this finding highlights that a high disease burden alone may not sufficiently motivate individuals to be vaccinated [24], especially in a context with the widespread belief that African countries are less susceptible to COVID-19 [25].

Our study highlighted some predictors of acceptance of COVID-19 vaccination.

### Healthcare workers and the general population

Adulthood was positively associated with the acceptance of the COVID-19 vaccine. A similar finding was found in previous studies [22] and is encouraging, given that these individuals are at a higher risk of being infected with SARS-CoV-2 [22]. Having a good vaccination intention did not guarantee the acceptance of a COVID-19 vaccine. A similar observation has been made elsewhere [26]. There are, therefore, obstacles in moving from the intention to vaccinate to actual vaccine intake [26]. Further research is needed to overcome these barriers.

Pregnant women were less likely to be vaccinated than those who were not pregnant. In this sense, a systematic review and meta-analysis showed that overall acceptance of the COVID-19 vaccine was low in pregnant women [27]. And would likely be due to the limited evidence on the safety of vaccines during pregnancy early in the pandemic and the conflicting and changing advice given to pregnant women as the pandemic evolved [28].

Acceptance of vaccination was low among healthcare workers who knew about vaccination compared with those who did not know. One possible reason is greater hesitancy among people with scientific information about vaccines’ potential side effects and risks [29]. The publication of scientific data on the safety of COVID-19 vaccines could increase their acceptance by healthcare providers [30].

In contrast, in the general population, having knowledge about vaccination was positively associated with vaccine acceptance. In this sense, strengthening knowledge about COVID-19 should be an effective way to increase the willingness to vaccinate [25]. Social norms are a significant predictor of health behaviours [31]. They can guide or constrain adopting a specific behaviour [32]. In our study, the likelihood of vaccination was greater among participants with favourable subjective norms.

### HCWs

Married women were more likely to be vaccinated than single women. Similar results have been found in other studies [26,33]. Married women would have a sense of more extraordinary family and collective responsibility [34]. The acceptance of a COVID-19 vaccine differed depending on the level of education. Professionally educated women were more likely to accept vaccination than those in high school. There was greater acceptance of vaccination as the years of schooling increased [34,35]. However, midwives showed more significant approval of vaccines compared to nurse assistants. Vaccine hesitancy has been reported among nurses [36,37]. This situation is concerning because patients are often in contact with patients and are frequently responsible for administering vaccines directly [36].

### GP

Acceptance of a COVID-19 vaccine was better in hypertensive women, which was encouraging. Indeed, a nearly 2.5-fold increase in the risk of severe COVID-19 in hypertensive patients has been reported to have an equally significant higher risk of mortality [38]. Also, we have seen greater acceptance of the COVID-19 vaccine among overweight/obese people. These individuals may be at a greater risk of experiencing the severe consequences of COVID-19 (hospitalisation, intensive clinical care and death) [42]. A positive perception of vaccination and the opportunity to be vaccinated promoted participants’ acceptance of the COVID-19 vaccine. Indeed, people who perceive vaccination as crucial in maintaining health are more likely to accept vaccines [43].

### Strengths and limitations

One of the first to combine the health workforce and the general population is identifying subgroups requiring attention. However, it was subject to certain limitations that are worth mentioning. First, it was a cross-sectional study and, therefore, unable to establish causality because of its cross-sectionality design. Second, its generalisability is limited only to persons active in the workplace and not to household members. We considered that people in the workplace were more exposed to risk because they interacted with others. Third, we have dichotomised the scales of some variables; this results in a loss of information but produces a more candid picture to classify behaviours.

### Conclusion

Our study showed a higher vaccination rate among healthcare workers than the general population. Favourable subjective norms and the ability to receive the vaccine have facilitated the acceptance of COVID-19 vaccination. Good vaccination intention was not sufficient for vaccine adoption. Youth, pregnancy and low educational attainment have been barriers to approving a COVID-19 vaccine. Exploring barriers and facilitators of immunisation helped identify these vulnerable subgroups that require attention. These findings could help design effective vaccination strategies to increase the acceptance of COVID-19 vaccines.

## Data Availability

The dataset and R code supporting this article's conclusion are included in the article (supplementary material).

## Conflicts of interest

The authors state that no conflicts exist between the parties cited in this study.

## Funding Statement

The National Centre for Training and Research in Rural Health of Maferinyah supported the field survey of this study under grant number 002/CNFRSR/2021 through only AAT. The funder had not played any role in this study.

## Acknowledgements

We thank the Guinean health authorities for facilitating the investigation. We also thank the National Center for Training and Research in Rural Health of Mafèrinyah and the Kofi Annan University for their financial and administrative support.

## Authors’ Contribution

Conceptualisation: Almamy Amara Touré, Ibrahima Barry.

Data curation: Almamy Amara Touré, Aboubacar Sidiki Magassouba, Younoussa Sylla.

Formal analysis: Almamy Amara Touré, Aboubacar Sidiki Magassouba.

Investigation: Kadiatou Bah, Alsény Yarie CAMARA.

Methodology: Almamy Amara Touré, Ibrahima Barry, Diao Cissé.

Resources: Almamy Amara Touré, Ibrahima Barry.

Software: Almamy Amara Touré.

Supervision : Abdourahamane DIALLO, Gaspard LOUA

Validation: Almamy Amara Touré, Ibrahima Barry.

Funding acquisition: Almamy Amara Touré.

Writing–original draft: Almamy Amara Touré, Ibrahima Barry

Writing–review & editing: Almamy Amara Touré, Ibrahima Barry, Aboubacar Sidiki Magassouba, Abdourahamane DIALLO

## Supplementary Material

Supplementary 1: additional_File1 Healthcare_workers

Supplementary 2: additionnal_File2 Healthcare_workers

Supplementary 3: additional_File3 General_Population.v1

Supplementary 4: additionnal_File4 General_Population

Supplementary 5: R code

## Notes

### Competing Interest Statement

The authors have declared no competing interest.

